# Structural Similarity Networks Reveal Brain Vulnerability in Dementia

**DOI:** 10.1101/2025.06.10.25328978

**Authors:** Marcella Montagnese, Amir Ebneabbasi, Natalia García-San-Martín, Clara Pecci-Terroba, Rafael Romero-García, Sarah E. Morgan, James H. Cole, Jakob Seidlitz, Timothy Rittman, Richard A.I. Bethlehem

**Affiliations:** Department of Psychology, University of Cambridge, Downing Pl, Cambridge, CB2 3EB, UK; Department of Clinical Neurosciences, University of Cambridge, Herchel Smith Building, Robinson Way, Cambridge, CB2 0SZ, UK; Department of Medical Physiology and Biophysics, University of Seville, C. San Fernando, 4, 41004 Sevilla, Spain; Department of Psychiatry, University of Cambridge, Herchel Smith Building, Robinson Way, Cambridge, CB2 0SZ, UK; Instituto de Biomedicina de Sevilla (IBiS), HUVR/CSIC/University of Seville / CIBERSAM, ISCIII, C. Antonio Maura Montaner, 41013 Sevilla, Spain; School of Biomedical Engineering and Imaging Sciences, King’s College London, Becket House, 1 Lambeth Palace Road, London SE1 7EU, UK; Department of Computer Science and Technology, University of Cambridge, William Gates Building, 15 JJ Thomson Ave, Cambridge CB3 0FD, UK; Department of Computer Science, Hawkes Institute, University College London, 90 High Holborn, London WC1V 6LJ, UK; Dementia Research Centre, UCL Queen Square Institute of Neurology, Queen Square, London, WC1N 3BG, UK; Lifespan Brain Institute, The Children’s Hospital of Philadelphia and Penn Medicine, 3401 Civic Center Blvd, Philadelphia, PA, 19104, USA; Department of Psychiatry, University of Pennsylvania, Philadelphia, PA 19104, USA; Department of Child and Adolescent Psychiatry and Behavioral Science, The Children’s Hospital of Philadelphia, 3401 Civic Center Blvd, Philadelphia, PA 19104, USA; Institute for Translational Medicine and Therapeutics, University of Pennsylvania, 3400 Civic Center Boulevard, Building 421, Philadelphia, PA 19104-5158, USA; Autism Research Centre, Department of Psychiatry, University of Cambridge, Douglas House, 18b Trumpington Road, Cambridge, CB2 8AH, UK; Brain Mapping Unit, Department of Psychiatry, University of Cambridge, Forvie Site, Robinson Way, Cambridge, CB2 2PZ, UK

**Keywords:** Normative modelling, MIND networks, Alzheimer’s disease, personalised medicine, neurodegeneration, structural MRI, neuropathology, brain networks, morphometric similarity

## Abstract

**INTRODUCTION:** Alzheimer’s disease (AD) is characterised by inter-individual heterogeneity in brain degeneration, limiting diagnostic and prognostic precision. We present a novel framework integrating Morphometric Inverse Divergence (MIND) networks with hierarchical Bayesian large-scale population modelling to identify individual-level neuroanatomical deviations.

**METHODS:** MIND networks quantify similarity between brain regions using multivariate MRI features. A normative model of regional MIND values trained on UK Biobank (N=35,133) was applied to the National Alzheimer’s Coordinating Center cohort (N=3,567). We examined brain deviations across clinical stages, APOE genotypes, mortality risk, and neuropathological burden.

**RESULTS:** Negative deviations (reduced MIND) stratified disease stages (p<0.01) and showed functional network enrichment in AD. Greater negative deviations characterised APOE ε4 homozygotes and correlated with postmortem neuropathological severity (p=0.032). Neurobiological decoding revealed associations with neurotransmitter receptor distributions and cortical organisation properties.

**DISCUSSION:** This population neuroimaging modelling enables individualised brain mapping with direct utility for diagnosis, prognosis, and understanding of biological mechanisms.

## 1. Background

Alzheimer’s disease (AD) represents a complex neurodegenerative syndrome characterised by heterogeneity in clinical presentation, progression, and underlying biological mechanisms^1,2^. Despite progress, prevailing neuroimaging approaches continue to rely on group-level comparisons—typically contrasting AD patients with cognitively healthy controls—potentially obscuring meaningful individual differences and limiting precision in diagnosis, prognosis, and treatment^3^. As personalised medicine gains traction across neurology and psychiatry, there is a pressing need for imaging frameworks that can provide reliable, individual-level characterisations of neurodegeneration.

Most neuroimaging studies rely on univariate approaches, examining features like cortical thickness or volume one at a time. While informative, such regionally specific approaches overlook the integrated, multivariate nature of cortical organisation^4^. The brain operates as an interconnected system, and growing evidence demonstrates that structural relationships between regions provide insights beyond individual regional properties^4–6^. This integrative approach may therefore offer crucial insights into dementia-related brain patterns that are missed by traditional approaches. Recent advances enable the integration of multiple features into multivariate similarity networks. Among these, Morphometric INverse Divergence (MIND)^7^ offers a biologically grounded method for constructing structural brain networks across cortical regions for each person, using more routinely collected structural T1-weighted MRI, enhancing their translational potential. MIND quantifies how similar each brain region is to all others by estimating similarity between vertex-level feature distributions across different quantitative morphometric measures like thickness, curvature, volume. The resulting MIND degree provides a summary measure of how structurally integrated a region is across the cortex, capturing subtle relationships that single-feature regional analyses may overlook. It also provides a biologically grounded representation of brain architecture that is more closely aligned with cortical cytoarchitectonic patterns and has stronger correspondence to axonal tract-tracing data than earlier approaches^7^.

Interpreting these complex networks at the individual level requires a statistical framework for distinguishing pathological change from normal variation and healthy ageing. Neuroanatomical population modelling provides such a framework by quantifying how much an individual deviates from expected patterns based on a healthy reference population^8–10^. Rather than comparing patients and controls at the group level, population modelling quantifies how each individual’s brain deviates from expected patterns for their demographic profile, with most advanced implementations accounting for confounding factors such as age, sex, and site/scanner effects. This approach creates personalised benchmarks that highlight complex brain patterns unique to each patient, while univariate approaches provide more focused deviations in specific measures. This method has successfully provided individual-level insights into neurodevelopmental and psychiatric disorders^11–17^. While population modelling is well established in these fields, its application to neurodegenerative disease is more recent but rapidly expanding. Important recent normative work in dementia^18,19^ has successfully applied this approach to cortical thickness and regional volume, demonstrating associations with cognitive decline and diagnostic conversion over time^18–21^. This work has provided useful insights into disease progression and heterogeneity cross-sectionally and longitudinally^18,20^, focusing on univariate models. Multivariate, network-based approaches remain largely unexplored in large-scale clinical datasets.

Building on this foundation, the National Alzheimer’s Coordinating Center (NACC) database^22^ offers a vital opportunity to advance a hybrid normative modelling and multivariate morphometric approach in a clinical context— providing clinical, imaging, genetic, and neuropathological data for robust analyses of brain deviations.

Our study extends previous work by leveraging multivariate MIND networks in the Alzheimer’s Disease Dementia spectrum for the first time. Critically, we introduce a novel integration of MIND with hierarchical Bayesian normative modelling to estimate individualised brain deviation maps. We apply this framework in a large, clinically heterogeneous cohort (NACC) to examine how individualised deviations relate to diagnostic status, APOE ε4 genotype, survival, and neuropathological markers from postmortem data. In doing so, we examine the multiscale complexity of dementia and its marked inter-individual variability. Finally, we explore whether spatial deviation patterns reflect biologically organised systems, including cortical hierarchy, neurotransmitters density, and metabolism, aiming to uncover structural signatures of disease that are both clinically informative and biologically interpretable.

## 2. Methods

### 2.1 Participants

Data for this study were derived from two datasets: (i) a reference (training) dataset including healthy individuals from the UK Biobank employed for normative modelling, and (ii) a clinical dataset that included people with Alzheimer’s Disease (AD), Mild Cognitive Impairment (MCI), and age-matched cognitively healthy controls. The reference dataset from the UK Biobank^23^ included N=35,133 healthy participants between 44 and 81 years old (mean age 63.56 ± 7.55), with 53% females and 46.9% males. This dataset has been extensively described in prior work^23,24^ and was used to train the initial normative model. Further details about the demographics and model implementation for this dataset can be found in **Supplementary Material Section 1** and **Supplementary Figures 1-6**. The clinical test dataset was obtained from the National Alzheimer’s Coordinating Center (NACC) database^22^, a publicly accessible multisite cohort (involving multiple Alzheimer’s Disease Research Centers - ADRCs) that includes a wide range of clinical, demographic, neuropsychological, and neuropathological information. As presented in **Table 1**, the final samples used for this study included healthy controls (NC, total *N* = 2,018), patients diagnosed with Alzheimer’s Disease (AD, *N* = 618), and individuals with Mild Cognitive Impairment (MCI, total *N* = 931). All clinical diagnostic labels had already been provided for each patient in the NACC database. The MCI group was further subdivided into two categories based on cognitive trajectory: (1) MCI Stable (*N*= 632); and (2) MCI Progressive (*N*= 299), for those who showed cognitive decline, as measured as changes over five years from baseline scanning in the CDR^®^ Dementia Staging Instrument (Clinical Dementia Rating). Data for this study were obtained from for UDS visits conducted between September 2005 and December 2022.

**Table 1.** Final Samples for NACC Dataset-Showing Demographics and Clinical Characteristics by Diagnostic Group.

| Diagnostic Group | Age (Mean ± SD) | Gender (%) | Education (Mean ± SD) | MMSE (Mean ± SD) | CDR® (Mean ± SD) | MOCA (Mean ± SD) |
| --- | --- | --- | --- | --- | --- | --- |
| **AD (n=618)** | 73.32±9.63 | F: 330 (53.40%),<br>M: 288 (46.60%) | 14.58±3.55 | 20.45±5.86<sup>^</sup> | 0.99±0.60 | 14.29±6.14<sup>^</sup> |
| **MCI Stable (n=632)** | 71.55±9.09 | F: 350 (55.38%),<br>M: 282 (44.62%) | 15.10±3.53<sup>^</sup> | 27.06±2.54<sup>^</sup> | 0.44±0.19 | 23.09±3.34<sup>^</sup> |
| **MCI Progressive (n=299)** | 74.56±7.42 | F: 153 (51.17%),<br>M: 146 (48.83%) | 15.34±3.32 | 26.02±2.67<sup>^</sup> | 0.51±0.13 | 20.67±3.44<sup>^</sup> |
| **NC<sub>test</sub> (N=394)** | 66.11±12.90 | F: 279 (70.81%),<br>M: 115 (29.19%) | 16.04±2.66<sup>^</sup> | 28.96±1.27<sup>^</sup> | 0.07±0.17 | 26.61±2.50<sup>^</sup> |
| **NC<sub>Train</sub> (N=1624)** | 66.19±10.57 | F: 1121 (69.03%),<br>M: 503 (30.97%) | 15.96±2.87 | 28.97±1.33<sup>^</sup> | 0.07±0.17 | 26.72±2.52<sup>^</sup> |

<sup>^</sup> Indicates missing values

#### 2.1.1 NACC Participant Characteristics and Cognitive Measures

Key demographic and clinical variables were obtained from the National Alzheimer’s Coordinating Center Uniform Data Set (UDS). Years of education reflect the total years of formal schooling as recorded in the UDS. APOE genotype was derived from available genetic data and reflects allele frequencies for ε2, ε3, and ε4, as provided by NACC. Cognitive function was indexed using the Mini-Mental State Examination (MMSE) and the Montreal Cognitive Assessment (MoCA), and global disease severity was captured using the CDR^®^ Dementia Staging Instrument. All values represent those recorded at the participants’ initial visit and are summarised in **Table 1** below.

### 2.2 MRI Acquisition and Preprocessing

Detailed MRI protocols, including T1-weighted sequences, are available online for both the clinical dataset ^22^ and the UK Biobank reference dataset^24^. For the reference cohort FreeSurfer version 6.0.1 was used for preprocessing, while FreeSurfer version 7.2 was used for NACC. For quality controls, following the methodology outlined in previous studies^8^, we excluded scans with a Euler Index (which quantifies topological defects/holes in the surface reconstruction of both hemispheres) of over N=2 Median Absolute Deviations (MAD). Additionally, to address potential confounding by head size, outliers in estimated Total Intracranial Volume (eTIV) we removed, defined as values outside the interquartile range for each diagnostic and sex group. Detailed exclusion numbers and proportions are provided in **Supplementary Section 1**. The final sample composition is detailed in **Table 1** and our analyses included data from 13 Alzheimer’s Disease Research Centers (ADRCs) in the adaptation and test set.

2.2.1 Morphometric INverse Divergence (MIND) Networks Construction and degrees estimation Morphometric INverse Divergence (MIND) networks were computed to characterise individualised structural similarity across the cortex, and have been described in detail in previous work^7^. Briefly, this approach estimates pairwise anatomical similarity between brain regions by quantifying divergence in multiple morphometric features derived from T1-weighted MRI. In our study, we extracted five regional features using FreeSurfer’s *recon-all* pipeline, namely: cortical thickness, mean curvature, sulcal depth, surface area, and grey matter volume. These features were chosen in accordance with prior studies^7,25^. For each pair of cortical regions, we calculated the symmetric Kullback–Leibler divergence between their multivariate feature distributions, resulting in a 360 × 360 symmetric matrix (based on the Human Connectome Project Multimodal (HCP-MMP) Glasser parcellation^26^; values bounded between 0 and 1) representing inverse divergence values. Higher values in this matrix reflect greater anatomical similarity. These matrices were used to generate individualised structural similarity networks, where each node corresponds to a cortical region, and edges represent the morphometric similarity between pair of regions. We then computed the weighted degree of each node by averaging its connectivity with all other regions, producing a 360-dimensional vector of regional similarity scores per subject (one score per cortical region in our parcellation). Importantly, here weighted MIND degree reflects the average similarity (in its multivariate morphometric profile) between a region and all others^7^, which can be thought of as a proxy for how morphometrically integrated it is. Detailed quality control analyses of feature contributions and network construction characteristics are provided in **Supplementary Figure 7.**

#### 2.2.2 Normative modelling of MIND Networks

We first trained a normative model on MIND weighted degree (derived as outlined in the previous section) from T1-weighted MRI data of *N*=35,133 healthy UK Biobank participants. We employed a hierarchical Bayesian regression (HBR) model^27^ due to its robustness in handling multisite datasets^28^, allowing for the integration of site-specific variability while accounting for confounding factors such as scanner differences and sampling biases. This method has proven particularly effective and has been employed in previous studies^18,29^. The model was implemented using the openly available *Predictive Clinical Neuroscience (PCN)* toolkit^30^ (https://github.com/predictive-clinical-neuroscience/braincharts) and employed a sinh-arcsinh (SHASHb) likelihood for enhanced modelling flexibility and to capture non-Gaussian characteristics in the data distributions^28^. Age and sex were included as fixed effects, while scanning site was modelled as a random effect. This approach appropriately models site differences as random offsets from a group mean while sharing information across sites. We implemented separate normative models for each of the 360 regions in our parcellation. Model performance evaluation including explained variance distributions across regions is shown in **Supplementary Figure 2**. All analyses were run on the Cambridge High-Performance Cluster.

Following initial training on the UK Biobank dataset, we employed an adaptive transfer learning approach^31^ to calibrate the estimates to our clinical dataset from NACC. We used 80% of the NACC healthy controls (NC_train_, *N*=1,624) to recalibrate the model parameters, establishing a NACC-specific normative baseline. The remaining 20% of healthy controls (NC_test_, *N*=394), along with all patients diagnosed with Alzheimer’s Disease (AD, *N*=618), MCI Stable (*N*=632), and MCI Progressive (*N*=299), formed the test set for evaluating deviations from the normative model (total test set *N*=1,943). All sites included in the test set were also represented in the adaptation dataset to ensure proper calibration of site-specific parameters. Further details on the specifics of the model can be found in the **Supplementary Section 1**.

For each participant in the test set, regional Z-scores were calculated after fitting a separate normative model for each region. These Z-scores quantify the degree of deviation from typical structural patterns observed in healthy aging. We defined extreme deviations as regions with Z-scores exceeding |1.96|, corresponding to a p-value of <0.05, following thresholds established in previous work^29^. Regions where Z-scores exceeded this threshold were considered as extreme deviations. Each participant’s total deviations count was computed as the sum of brain regions with such extreme deviations, again to stay in line with previous work^29^. Most analyses were performed separately on positive and negative deviations to better capture the direction-specific nature of brain changes, as these deviations may reflect different underlying neurodegeneration processes. To account for the potential confounding effect of head size, we residualised regional Z-scores against eTIV separately for each demographic group (by sex and diagnosis), ensuring findings reflect disease-related changes rather than anatomical variations due to head size (full details in **Supplementary Section 1** and **Supplementary Figures 5 and 6**). Quality control analyses confirmed model robustness, with no significant correlation between age and deviations (r = −0.026, p = 0.250). Further details on model implementation, parameters, and quality control can be found in **Supplementary Material Section 1.** All analyses were run in *Python 3.12.4*.

### 2.3 Group-Level Deviations and Network-Specific Targeting

To assess how MIND-based deviations stratify clinical stages and target known brain systems, we first compared deviations across all diagnostic groups. For each participant, positive and negative deviation counts — defined as the total number of cortical regions with Z-scores exceeding +1.96 or below –1.96, respectively — were calculated, corresponding to the top and bottom 2.5% of the normative distribution (two-tailed p < 0.05). Separate one-way ANOVAs were conducted to compare positive and negative total deviation counts across groups (AD, MCI Progressive, MCI Stable, and Healthy Controls (NC)). Post-hoc comparisons were performed using Tukey’s HSD test to identify pairwise group differences.

To examine spatial patterns of deviations, we implemented a regional overlap analysis based on work by Segal et al. (2023)^16^. For each region and diagnostic group, we computed the proportion of individuals with positive and negative deviations. We then subtracted the corresponding overlap rate in the held-out healthy control group (NC_test_) from each clinical group to create deviation difference maps, highlighting regional increases or decreases in deviation prevalence. To assess statistical significance, we used permutation testing (*N* = 10,000), shuffling diagnostic labels to generate null distributions of regional overlap differences while preserving group sizes. Directional p-values were computed for each region (both patient > control and control > patient), and false discovery rate (FDR) correction was applied.

We next tested whether deviations aggregated within known brain systems using a network-wise overlap analysis, based once again on previously published work in psychiatric cohorts by Segal and colleagues^16^. For each of the Yeo and Krienen seven functional networks^32^, we computed the proportion of individuals in each group with at least one extreme deviation in the network (|Z| > 1.96). Normative benchmarks were computed from held-out NC participants. Δ-overlap maps were derived by subtracting this control overlap map from each clinical group’s map.

To statistically assess network-level enrichment, we implemented two complementary null models. First, we used group-label permutation testing (*N* = 10,000), which preserved group size but randomly reassigned diagnostic labels. Second, to ensure observed network differences could not be explained by chance spatial structure or global deviation burden, we generated spatially constrained null maps using the Hungarian algorithm with the *neuromaps* toolbox^33^, preserving the spatial structure of cortical maps while randomizing spatial location. Both nulls were used to generate empirical p-values, with FDR correction applied across networks.

Lastly, to complement this categorical network-level approach with a continuous group-level contrast, we performed permutation-based significance testing of regional Cohen’s d effect sizes across three pairwise diagnostic comparisons: MCI Stable versus MCI Progressive, MCI Stable versus AD, and MCI Progressive versus AD. For each comparison, we calculated observed Cohen’s d values between groups for each ROI’s deviation scores, with effect directionality set such that the more severe diagnostic group was always subtracted from the less severe group to aid interpretability. Statistical significance was determined through *N*=10,000 random permutations of diagnostic labels, generating empirical two-tailed p-values, with FDR correction applied to address multiple comparisons. These effect size maps quantified structural deviations across key stages of clinical progression within the Glasser atlas parcellation. While Cohen’s d comparisons highlight robust group-level effects, they assume within-group homogeneity. To assess this assumption, we computed pairwise Hamming distances between individual binary deviation maps for negative and positive deviations separately.

### 2.4 Genetic Stratification by APOE Genotype

We next explored whether deviations reflect underlying genetic risk. In line with established APOE classifications used in recent work^34^, participants were grouped into four APOE genotype categories: ε3 homozygotes (*reference* group), ε2 carriers (ε2/ε2, ε2/ε3), ε4 heterozygotes (ε3/ε4), and ε4 homozygotes (ε4/ε4). Individuals with ε2/ε4 genotypes were excluded due to opposing risk profiles, and those missing APOE data were removed^34^. Final group sizes were: ε3 homozygotes (*N* = 556), ε4 heterozygotes (*N* = 517), ε4 homozygotes (*N* = 133), and ε2 carriers (*N* = 93).

We analysed two deviation metrics derived from MIND normative modelling: (1) positive deviation count (number of regions with z > 1.96), and (2) negative deviation count (number of regions with z < –1.96). For each outcome, we fitted a series of nested ordinary least squares (OLS) regression models with APOE group as a categorical predictor (ε3 homozygotes as reference) and age and sex as covariates. To assess the presence of interactions, we systematically tested: (1) a basic additive model; (2) models including APOE × age interactions; (3) models including APOE × sex interactions; and (4) a full model with all two-way and three-way interactions. Model selection was performed using Akaike Information Criterion (AIC), with likelihood ratio tests used to evaluate the significance of interaction terms against the basic model. Post-hoc pairwise comparisons between APOE groups were conducted using Tukey’s test.

### 2.5 Prognostic Analysis: Mortality

To evaluate whether brain deviations predicted mortality we used Cox proportional hazards regression. Consistent with prior work of normative modelling in Alzheimer’s disease^20,35^, for this analysis we focused on negative deviations only, which more reliably reflect clinical progression. Survival time was defined as the interval (in years) between age at MRI acquisition and either age at death (for deceased individuals) or age at last follow-up (for censored individuals). A total of N=558 participants died during follow-up (AD: 330, MCI Progressive: 134, MCI Stable: 94), while N=991 participants (AD: 288, MCI Progressive: 165, MCI Stable: 538) were right-censored at their last known follow-up, determined from longitudinal NACC dataset records.

We used the previously defined negative deviation count (number of cortical regions with z-scores < –1.96) as a continuous predictor in the survival model. A Cox proportional hazards model was initially fit including age at MRI, sex, negative deviation count, and diagnostic group as covariates. Proportional hazards assumptions were assessed using scaled Schoenfeld residuals. While individual covariates (age, sex, negative deviation count) met the proportional hazards assumption (all p > 0.05), a violation was observed for the diagnostic group variable (p < 0.001). To address this violation, we employed two complementary approaches: i) a stratified Cox model by diagnostic group that allowed different baseline hazard functions while estimating a common effect of deviation count across groups, and ii) Cox models fitted separately for each diagnostic group to evaluate within-group effects and assess potential heterogeneity in prognostic value across disease stages.

For visualization purposes, the total count of cortical regions with z-scores below –1.96 was binarized at the cohort median to create a high vs. low deviation group. Kaplan–Meier survival curves were plotted using this binary classification, with survival differences evaluated using log-rank tests. All survival analyses were conducted in *R version 4.5.0*.

### 2.6 Neurobiological Decoding of Structural Deviations

To enhance mechanistic interpretations of structural deviations identified through normative modelling, we performed spatial co-location analyses between deviation maps and a comprehensive set of cortical reference maps. The analysis was done using the regional Z-scores for each diagnostic group (left hemisphere). Biological reference maps were drawn from established open-access repositories and covered a wide range of cortical properties. These included neurotransmitter receptor density distributions derived from PET meta-analyses (e.g., for serotonin 5-HT2a, dopamine D1, GABAa, and nicotinic α4β2 receptors —originally compiled by Hansen et al. (2022)^36^). For cytoarchitectonic features, we used information available from the BigBrain Atlas^37^ alongside available maps used in previous work^38–40^ that provided information about cortical metabolism (cerebral blood flow, glucose metabolism), microstructural proxies such as myelin (T1w/T2w), cortical hierarchy (Mesulam), developmental/evolutionary expansion maps^41^, and for transcriptomic the first principal component from the Allen Human Brain Atlas^42^. All these maps were retrieved from prior original work^36–41,43^ or from the *neuromaps* toolbox^33^. Please refer to the original sources for more detailed descriptions. To account for spatial autocorrelation when evaluating cortical map correlations, we applied spin permutation testing using the Hungarian rotation method. Cortical centroid coordinates were rotated on a spherical surface to generate N=5,000 spatially permuted versions of the parcellation. These permutations were then used to reorder the deviation maps, producing null distributions of correlation values for each biological reference map. Empirical spatial correlations were compared against these null distributions to derive spin-corrected *p***-**values, which were used to assess statistical significance.

### 2.7 Relationship with Postmortem Pathology

For a subset of participants (*N*=240) with autopsy-confirmed pathology in NACC (see further details in **Supplementary Section 6** and **Supplementary Table 7**), we examined associations between total deviation count and a combined neuropathological score available in this cohort^44^. Neuropathological analyses were performed in accordance with the National Institute on Aging–Alzheimer’s Association (NIA-AA) guidelines^45,46^ for the assessment of Alzheimer’s disease neuropathologic change (ADNC). In line with previous work^47^, the primary measure used for stratification was the ABC score (NACC variable: “NPADNC”), a composite ordinal index ranging from 0=“No AD pathology” to 3= “High AD pathology”, and that integrates three key pathological processes associated with Alzheimer’s disease: amyloid-β (Aβ) deposition, tau neurofibrillary degeneration, and neuritic plaque burden. Specifically, the ABC score provided in the NACC dataset reflects a combination of Thal phase (“A” component), Braak stage (“B” component), and CERAD rating of neuritic plaque density (“C” component).

To assess whether the number of deviations—positive and negative, respectively—was associated with the severity of neuropathological burden, we modelled the relationship between deviation count and ABC score using ordinal logistic regression. Age at MRI, sex, and time to death were included as covariates in all models. A basic model adjusted for demographic and clinical variables provided an initial assessment. An age interaction model introduced interaction terms between ABC score and age at MRI to investigate age-dependent neuroimaging and cognitive performance effects. A third model, with time-to-death interaction, explored disease progression dynamics. Model comparison was based on the Akaike Information Criterion (AIC). The main analysis focused on the composite ABC score, with exploratory analyses of the individual A, B, and C components conducted and reported in **Supplementary Material Section 6**. Participants with missing or invalid pathology scores (i.e., values of 8, 9, or –4 in the NACC dataset) were excluded from all relevant analyses. All regression models were implemented using the *statsmodels* Python library. Post-hoc comparisons were performed using Tukey’s test to evaluate pairwise differences between ABC levels while controlling for multiple comparisons. A two-sided significance threshold of p < 0.05 was used throughout.

## 3. Results

### 3.1 Group Differences in Positive and Negative Deviation Counts

There were no significant group differences in positive deviation count (ANOVA: F(3,1939) = 0.59, *p* = 0.62). In contrast, negative deviation count differed significantly across groups (ANOVA: F(3,1939) = 24.39, *p* < 0.001). Post-hoc Tukey tests revealed that individuals with AD had significantly higher negative deviation counts compared to all other groups (AD vs. MCI Progressive: *p* = 0.001; AD vs. MCI Stable and Healthy Controls (NC): *p* < 0.001). MCI Progressive individuals also showed more negative deviations than NC (*p* = 0.049), but not significantly more than MCI Stable (see **Supplementary Table 1** for full ANOVA and post-hoc results).

### 3.2 Spatial Overlap and Significance of Individual Deviations

To assess whether cortical deviations aggregated non-randomly across diagnostic groups, we computed the proportion of individuals with extreme positive or negative deviations for each region, generating group-level overlap maps (**Figure 1A**). While deviation overlap was generally low across the cortex, AD exhibited notably higher rates of overlap, particularly for negative deviations in medial temporal, lateral temporal, inferior parietal, and posterior cingulate cortices. Positive deviations were more spatially restricted and mostly in visual and ventral temporal regions.

**Figure 1.**
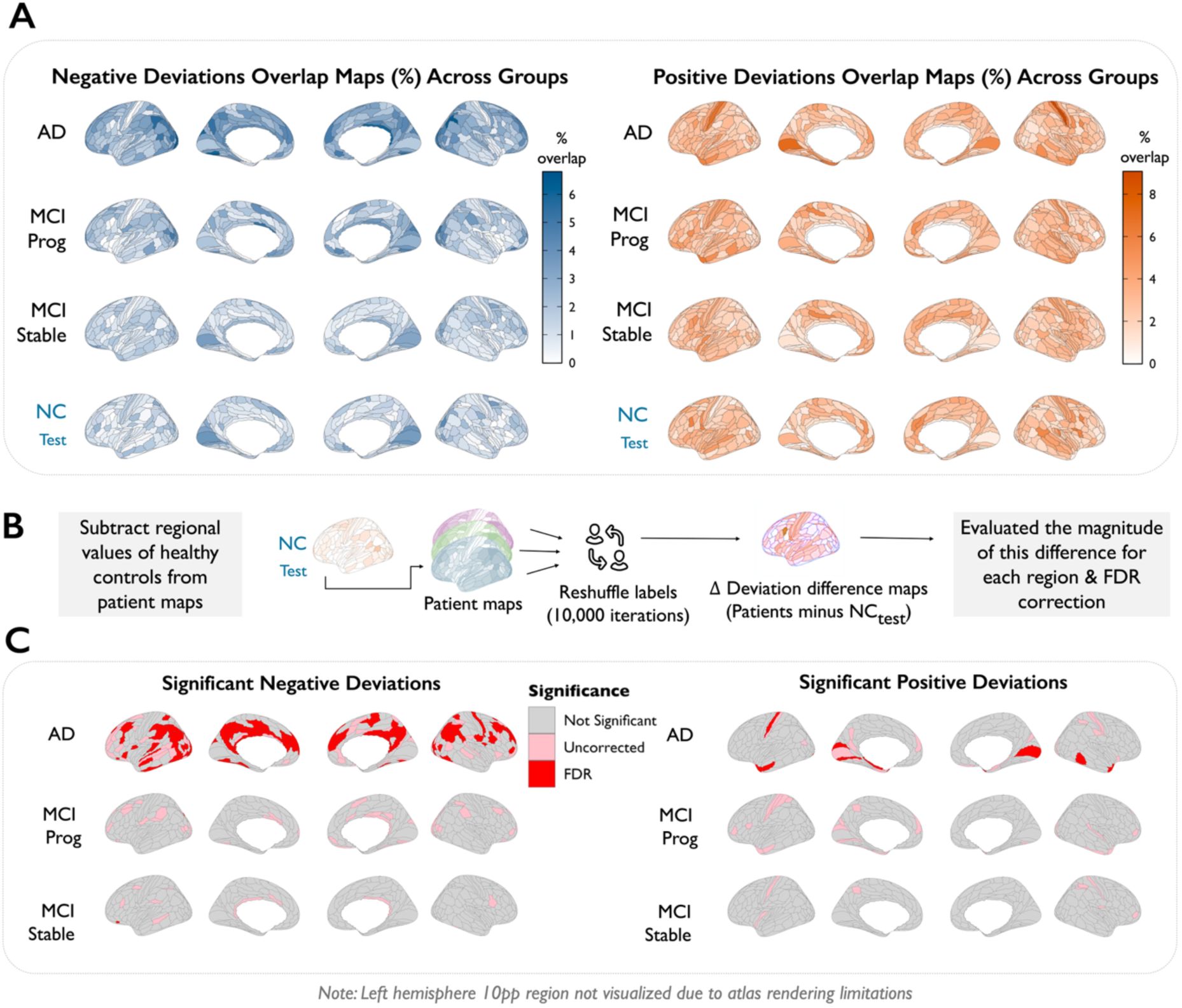
Group-level spatial overlap and statistical significance of cortical deviations. (**A**) Left in blue, regional overlap maps show the percentage of individuals within each diagnostic group exhibiting extreme negative deviations (Z < −1.96). Right in orange, regional overlap maps show the percentage of individuals within each diagnostic group exhibiting extreme positive deviations (Z > 1.96). (**B**) Schematic diagram explaining how we calculated the deviation difference maps **(C)** Left: Cortical regions with significantly higher negative deviation overlap relative to controls, based on permutation testing (N= 10,000). Right: Cortical regions with significantly higher positive deviation overlap relative to controls, based on permutation testing (N= 10,000). In both panels, red indicates regions significant after FDR correction, and pink shows uncorrected significance (p < 0.05). NC = normal controls (test set); MCI = mild cognitive impairment.

To determine whether observed patterns exceeded those expected by chance, we computed difference maps by subtracting control group overlap from each clinical group and used permutation testing to assess statistical significance (schematics shown in **Figure 1B**). Extensive clusters of FDR-significant negative deviations were observed in AD, with circumscribed effects in MCI Progressive, and only very few significant regions observed in MCI Stable although these did not survive FDR-correction (shown in lighter pink in left panel of **Figure 1C**). Positive deviations analysis showed fewer significant regions across all groups, with only some effects evident in AD remaining statistically significant after multiple comparison correction (Right Panel of **Figure 1C**).

### 3.3 Network-Level Enrichment of Cortical Deviations

We next assessed whether cortical deviations preferentially aggregated within each of the seven Yeo functional networks (**Figure 2 A-D**). Compared to controls, AD showed markedly elevated overlap for negative deviations in most networks, including the default mode network (DMN), both dorsal and ventral attention networks, frontoparietal network, and somatomotor network. The limbic network was significant for the group-nulls, but not for the spatial-null analyses, with the visual network not related to negative deviations in AD in either analysis. No significant differences were found when comparing either group of MCI patients to healthy controls for negative deviations.

**Figure 2.**
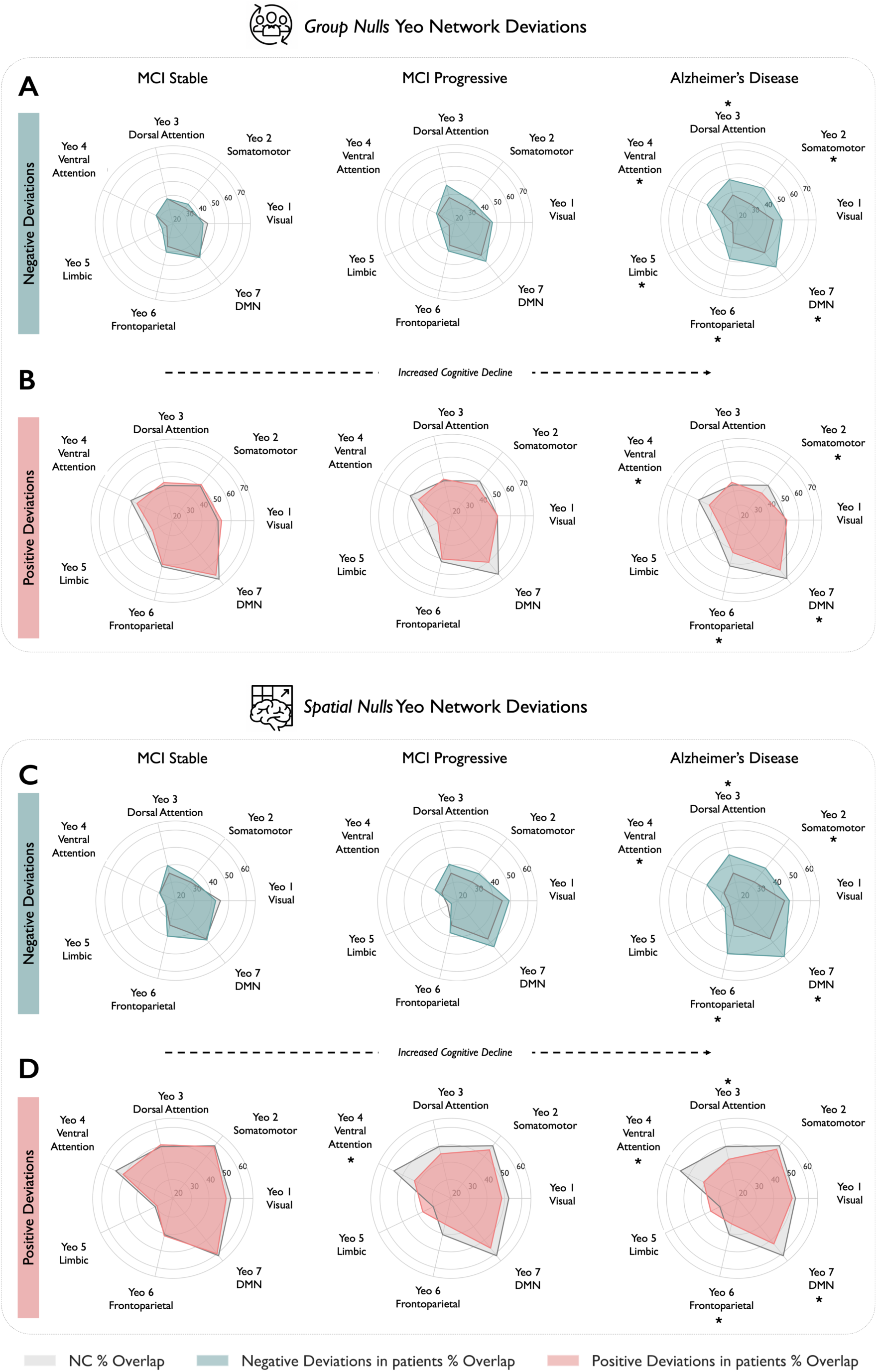
Network-level enrichment of cortical deviations in Alzheimer’s disease and prodromal stages. (A–B) Group-label permutation-based deviation overlap for negative (A) and positive (B) deviations across the Yeo 7-network parcellation. Radar plots display the percentage of individuals in each diagnostic group (MCI-Stable, MCI-Progressive, AD) exhibiting at least one extreme deviation (|Z| > 1.96) within each network. (C–D) Equivalent results using spatially constrained null models that preserve the spatial autocorrelation of cortical maps. Deviations are computed relative to a held-out healthy control benchmark (NC), with shaded regions representing differential overlap in patients versus controls. * indicates statistically significant differences (p < 0.05).

Positive deviations were more spatially diffuse, with both group- and spatial-null models converging to show a significantly diminished overlap in AD compared to controls in ventral attention, somatomotor, DMN, and frontoparietal network. Spatial null models also showed the same pattern for the ventral attention network in MCI Progressive.

#### Cohen’s d results

After FDR correction (full details in **Supplementary Table 2**), Cohen’s *d* effect sizes for deviations between key diagnostic comparisons showed that the largest effect sizes were observed, as expected, in the comparison between AD and MCI Stable (top row, **Figure 3**). For this comparison, widespread FDR-significant effects were observed across medial temporal regions (e.g., bilateral PHA2 in the parahippocampal gyrus) and retrosplenial and posterior cingulate areas (e.g., bilateral 23c, RSC). These disruptions extended into lateral and parietal cortices (e.g., POS2, PGi), ventrolateral prefrontal cortex (e.g., 47s, FOP3), and medial prefrontal regions (10v), closely tracking known hubs of the default mode and frontoparietal control networks. AD also showed significant disruption relative to MCI Progressive in overlapping areas (middle row, **Figure 3**), though with much more limited spatial extent (e.g., 23c, 24v and 47s in the left frontal lobe) and with the opposite direction of effect (i.e., suggesting less deviation in the AD than MCI Progressive), for the primary visual cortex (bilateral V1). Lastly, MCI Progressive individuals differed from the MCI Stable group only in a region of the dorsal margin of inferior parietal cortex (IPO), and in the perirhinal entorhinal cortex in the temporal lobe. Overall, these results reinforce the graded nature of deviation accumulation across clinical stages, with the most robust structural disruptions emerging in regions linked to memory, executive function, and the default mode network.

**Figure 3.**
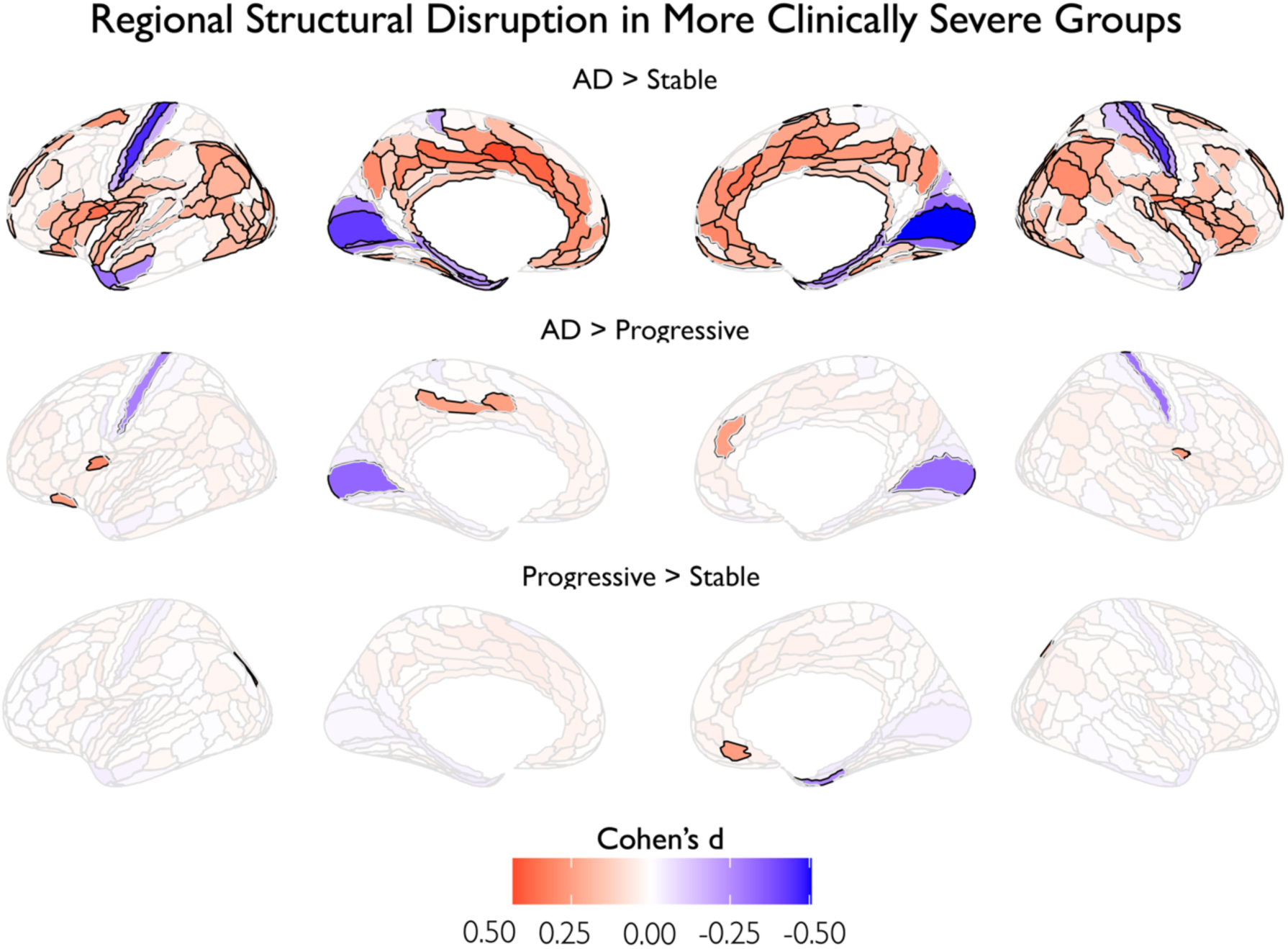
Cohen’s d maps show group differences in deviation from normative structure (Z-scored MIND-weighted degree). Maps are thresholded by FDR-corrected significance (black borders indicates significant P-val after FDR). Red indicates greater disruption in the more severe group; blue indicates greater disruption in the less severe group. Effect sizes reflect three pairwise comparisons: AD vs. MCI Stable, AD vs. MCI Progressive, and MCI Progressive vs. MCI Stable.

These group-level Cohen’s d results should be interpreted with caution, as they assume homogeneity within diagnostic groups—an assumption we directly tested using Hamming distance analysis, which revealed moderate inter-individual variability within each group. For negative deviations, median Hamming distances went from 7.0 in NC and MCI Stable, to 8.0 in MCI Progressive, and 11.0 in AD, with corresponding increases in interquartile ranges (NC and MCI Stable: IQR = 8.0; MCI Progressive: IQR = 10.0; AD: IQR = 16.0). For positive deviations, median Hamming distances were as follows: 13.0 in NC (IQR = 17.0), 12.0 in MCI Stable (IQR = 16.0), 12.0 in MCI Progressive (IQR = 17.0), and 12.0 in AD (IQR = 20.0).

### 3.4 APOE Analyses

Having established that MIND-based structural deviations are sensitive to disease stage, we next examined whether these deviations also reflect underlying genetic risk. Specifically, we tested for associations between APOE genotype and the number of cortical regions exhibiting extreme deviations. For positive deviations, the basic additive model showed the best fit (AIC = 11,130.3), with interaction models providing no improvement (likelihood ratio tests: age interaction p = 0.407, sex interaction p = 0.367). For negative deviations, the sex interaction model was the best one (AIC = 10,397.7), representing a significant improvement over the basic model (likelihood ratio test: p = 0.040). The full three-way interaction model (age*sex*deviations) showed no further improvements for either outcome (See **Supplementary Table 3**). The sex composition for the groups was as follows: ε3 homozygotes: N=289 females and N=267 males; ε2 carriers: N=39 females and N=54 males; ε4 heterozygotes: N=283 females and N=234 males; ε4 homozygotes: N=62 females and N=71 males.

For positive deviations, no interactions with age or sex were detected (See **Supplementary Table 4**). ε2 carriers exhibited significantly more positive deviations than ε3 homozygotes (β = 4.76, *p* = 0.016, **Figure 4A**). Neither ε4 heterozygotes (β = 0.74, *p* = 0.490) nor ε4 homozygotes (β = −1.26, *p* = 0.459) differed significantly from ε3 homozygotes.

**Figure 4.**
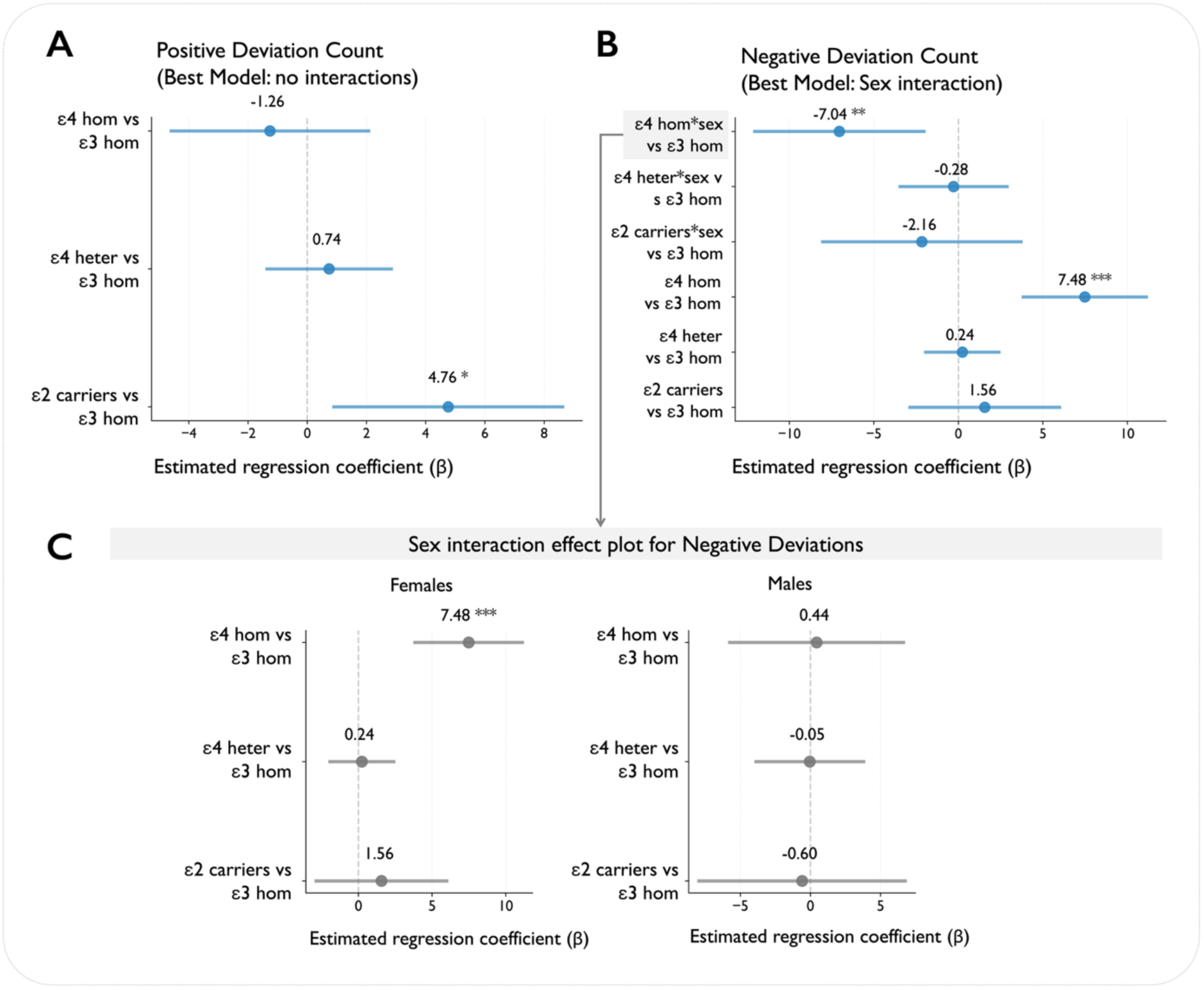
APOE genotype effects on brain structural deviation counts. **(A)** Forest plot displays regression coefficients (β ± 95% confidence intervals (CI)) for positive deviation count from the best-fitting model (no interactions). All estimates are relative to ε3 homozygotes and adjusted for age and sex. **(B)** Regression coefficients from the sex interaction model for negative deviation count, showing both main effects and interaction terms (APOE × sex). **(C)** Sex-stratified effects for negative deviations. Points represent effect estimates with 95%CI. Statistical significance: * p < 0.05, ** p < 0.01, *** p < 0.001.

For negative deviations, we observed a significant APOE*sex interaction (likelihood ratio test: p = 0.040), indicating sex-specific APOE effects. Overall, ε4 homozygotes showed substantially more negative deviations than ε3 homozygotes (**Figure 4B**), and this effect was predominantly observed in females (β = 7.48, *p* < 0.001), **Figure 4C**. In males, the ε4 homozygote effect was markedly attenuated (sex interaction term: β = −7.04, *p* = 0.006), resulting in a minimal overall effect in this group (β = 0.44). No age interactions were detected for negative deviations (p = 0.42). All details for these models and tests can be found in **Supplementary Tables 4 and 5**.

### 3.5 Negative Deviations Do Not Predict Mortality Risk in Alzheimer’s Disease

To assess whether individual structural brain deviations predicted survival, we ran two main analyses. The stratified Cox model revealed that continuous negative deviation count was not significantly associated with mortality risk (HR = 1.002, 95% CI: 0.997-1.007, p = 0.45). Separate Cox models for each diagnostic group (**Figure 5A)** confirmed that structural deviation burden was not predictive of mortality in any disease stage: AD (HR = 1.0005, 95% CI: 0.995-1.006, p = 0.86), MCI Progressive (HR = 1.008, 95% CI: 0.990-1.027, p = 0.40), and MCI Stable (HR = 1.008, 95% CI: 0.988-1.029, p = 0.43). Age at MRI showed the strongest prognostic value in MCI Stable patients (HR = 1.073, 95% CI: 1.044-1.102, p < 0.001), while showing minimal effect in AD (HR = 1.002, 95% CI: 0.989-1.016, p = 0.71) and MCI Progressive groups (HR = 1.013, 95% CI: 0.988-1.039, p = 0.31). Male sex remained a significant mortality predictor in AD patients (HR = 1.46, 95% CI: 1.17-1.81, p = 0.001) but not in MCI Progressive (HR = 1.24, 95% CI: 0.87-1.76, p = 0.23) or MCI Stable groups (HR = 1.42, 95% CI: 0.90-2.25, p = 0.11).

**Figure 5.**
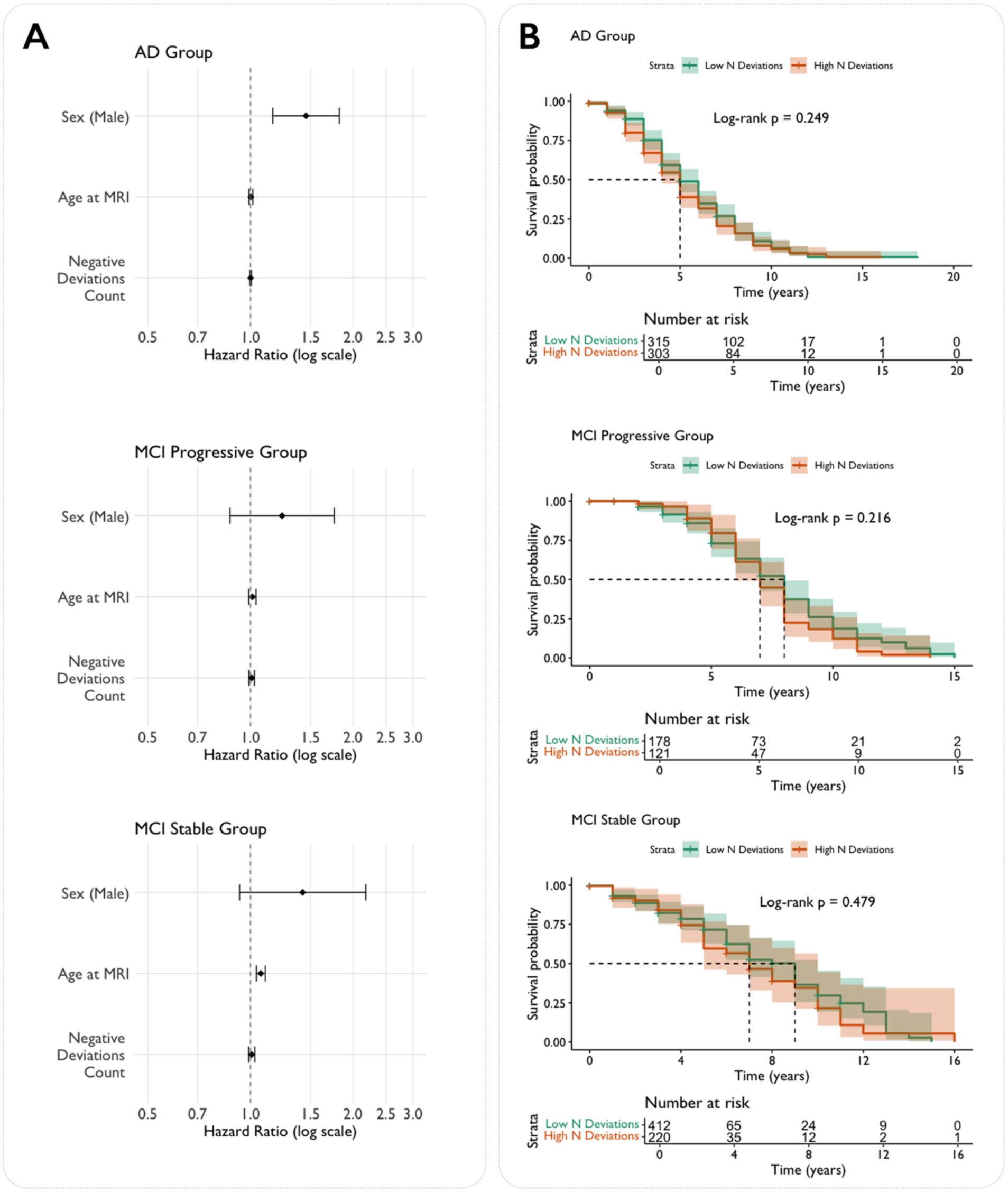
**(A)** Forest plots showing hazard ratios (HRs ± 95% confidence intervals (CI)) from separate Cox proportional hazards models for each diagnostic group, assessing the relationship between negative deviation count and survival. **(B)** Kaplan–Meier survival curves stratified by high vs. low structural deviation burden (median split within each diagnostic group) for AD, MCI Progressive, and MCI Stable groups. Shaded areas denote 95% CI. Log-rank p-values indicate no significant survival differences between high and low burden groups in any diagnostic category. * p < 0.05, ** p < 0.01, *** p < 0.001.

Kaplan–Meier survival curves stratified by high versus low structural deviation burden (divided at the cohort median) confirmed the Cox regression findings (**Figure 5B**). Log-rank tests revealed no statistically significant survival differences between high and low burden groups in any diagnostic category (all p > 0.05), consistent with the continuous analyses.

### 3.6 Neurobiological Decoding of normative MIND deviations

We assessed the spatial correspondence between regional normative deviations and a range of cortical biological properties across diagnostic groups (**Figure 6**). Full details can be found in **Supplementary Table 6**. When assessing different neurotransmitters, higher average deviations in the MCI Stable —characterised by predominantly positive Z-scores (see corresponding brain plot at the bottom left of **Figure 6**) —showed significant positive correlations with several receptor maps, including mGluR5 (*r* = 0.48, *p* <0.001), MOR (*r* = 0.48, *p* <0.001), GABAa (*r* = 0.39, *p* = 0.002), and different 5HT receptors. These associations were less strong in MCI Progressive and were reversed in AD, where deviations were mostly negative (**Figure 6**, bottom right). In fact, significant negative correlations were observed with 5HT2a (*r* = –0.35, *p* <0.001), 5HT4 (*r* = –0.31, *p* = 0.007), GABAa (*r* = –0.30, *p* = 0.005) and mGluR5 (*r* = –0.27, *p* =0.008), indicating that regions where MIND decreased compared to what expected from age- and sex-matched healthy population references, were partly spatially co-localised with areas of higher normative serotonergic, GABAergic and glutamatergic density.

**Figure 6.**
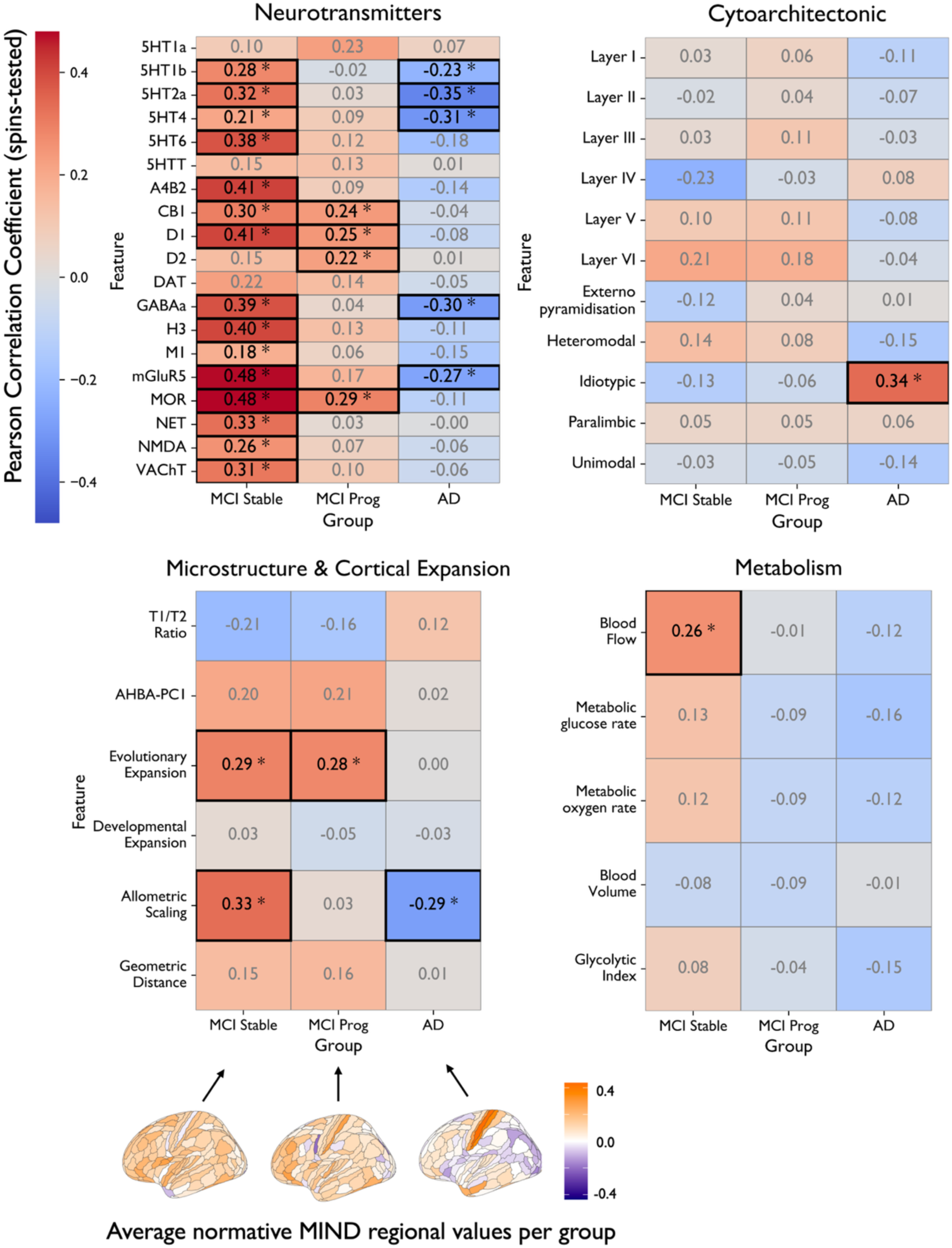
Biological decoding of group-average deviation maps. Heatmaps show Pearson correlation coefficients (r) between average regional MIND deviation maps (Z-scores) for each diagnostic group (columns: MCI Stable, MCI Progressive, AD) and a range of biological reference maps (rows), spanning four domains: neurotransmitter receptor density, cytoarchitecture, metabolism, and microstructure/ cortical expansion. Correlations were tested for statistical significance using spin permutation testing (N = 5000) to control for spatial autocorrelation; significant values (p < 0.05, spin-corrected) are marked with asterisks and outlined in black. Colour indicates the direction and magnitude of the correlation (red = positive, blue = negative). Group-average deviation maps are visualized below for reference with their corresponding legend. AHBA = Allen Human Brain Atlas. * indicates statistically significant differences (p < 0.05).

Average cortical deviations also aligned with structural expansion properties. MCI Stable and MCI Progressive groups, which showed predominantly positive deviation, demonstrated positive correlations with evolutionary expansion (*r* = 0.29 and 0.28; *p* = 0.005 and 0.014), alongside allometric scaling in the MCI Stable group (*r* = 0.33, *p* = 0.002), indicating that, on average, the more positive deviations occur in regions with higher evolutionary expansion characteristics and higher allometric scaling (MCI stable only). AD, characterised by predominantly negative deviations, showed the opposite direction of effect for allometric scaling (*r* = –0.29, *p* <0.001), potentially indicating that regions with higher allometric scaling showed greater negative deviations (greater structural disruption).

Among cytoarchitectonic features, only idiotypic cortex showed a significant association, with average cortical AD deviations being spatially co-localised with areas showing higher idiotypic characteristics (*r* = 0.34, *p* = 0.001). No other layers or classes survived correction.

Lastly, in the metabolic domain, only one feature reached significance: higher average normative deviations in MCI Stable were spatially co-localised with regions of higher blood flow (*r* = 0.26, *p* = 0.03), while no statistically significant results were observed in the MCI Progressive or AD groups.

### 3.7 Prognostic Analysis: More negative deviations are linked to higher post-mortem pathology

We investigated whether the burden of structural deviations was associated with neuropathological severity in a subset of N=240 participants for whom autopsy data were available in NACC (schematics shown in **Figure 7A**). Ordinal logistic regression revealed a significant association between the total number of extreme negative deviations and higher ABC score (**Figure 7B**). The best-fitting model included an interaction term with age at MRI (AIC = 427.8), indicating that the relationship between structural disruption and AD pathology was age-dependent. Specifically, the main effect of negative deviations was significant (*OR* = 1.96, 95% CI: 1.06–3.63, *p* = 0.032), with an additional negative interaction with age (*p* = 0.041). In contrast, the number of extreme positive deviations was not associated with ABC score in any models (all *p* > 0.1), and time-to-death interactions did not improve model fit. Details of all models and of the distribution of ABC scores across diagnostic categories can be found in **Supplementary Table 7** and **8**, and **Supplementary Figure 8**, with stratified model results shown in **Supplementary Table 9**.

**Figure 7.**
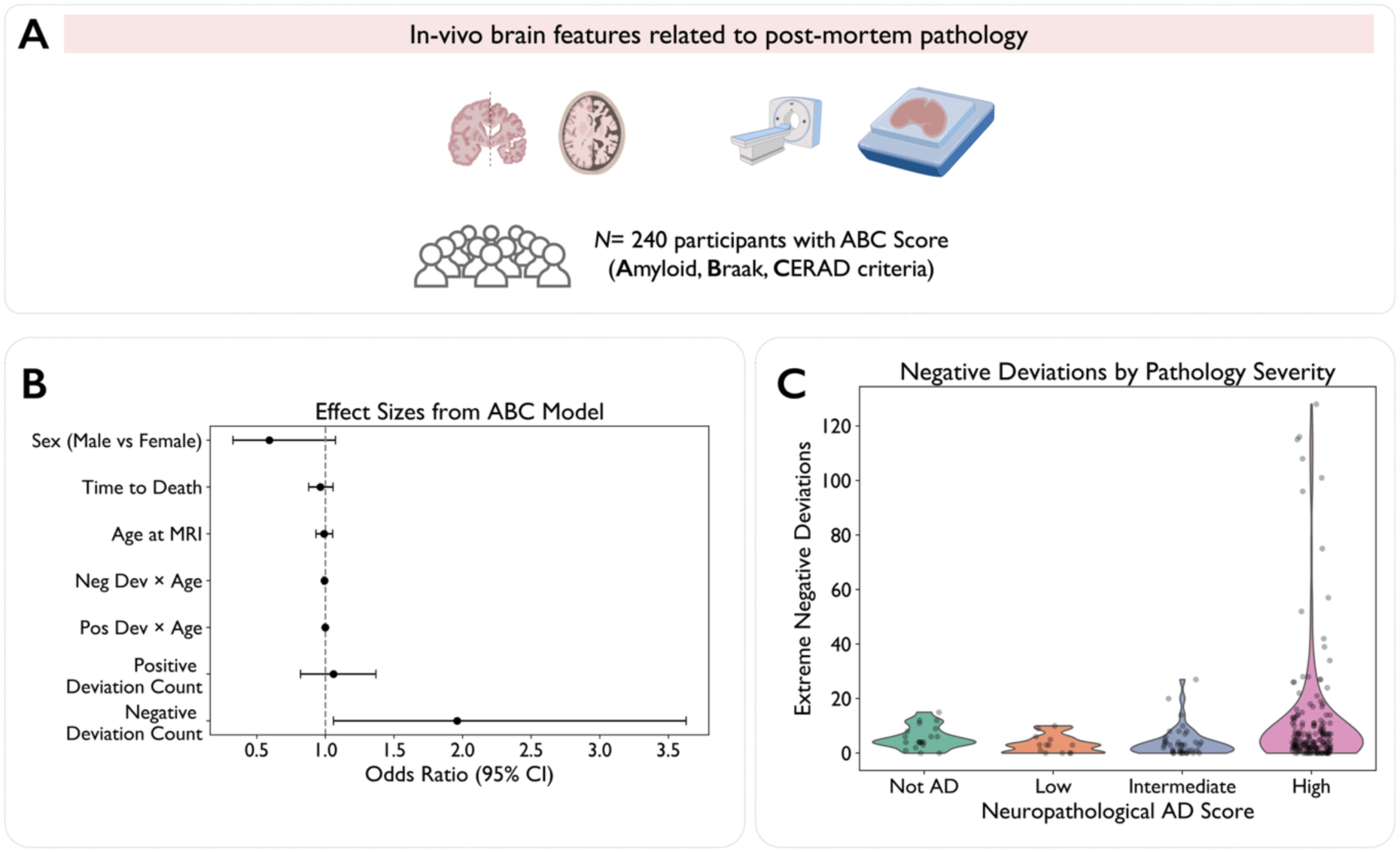
Structural deviation burden is associated with neuropathological severity. **(A)** Overview of the subsample (N = 240) from the National Alzheimer’s Coordinating Center (NACC) with in vivo MRI and subsequent ABC scoring (Amyloid, Braak, and CERAD criteria). **(B)** Forest plot showing odds ratios and 95% confidence intervals from the ordinal logistic regression model predicting ABC score**. (C)** Violin plots of extreme negative deviations across neuropathological severity levels (where neuropathological AD score refers to the ABC Scores).

Distribution of deviation burden across ABC pathology levels is shown in **Figure 7C**. Diagnostic stratification showed that this effect generalised across Alzheimer’s disease and progressive MCI, but was attenuated in MCI Stable participants, consistent with lower pathology burden in that group. Exploratory models examining each A, B, and C scores as outcomes did not reveal consistent associations with deviation count and are reported in **Supplementary Table 10**.

## 4. Discussion

### 4.1 Summary of Key Findings

This study presents two significant methodological advances: the first integration of MIND networks with hierarchical Bayesian normative modelling, and the first application of MIND to Alzheimer’s disease. These advances were leveraged to reveal individual-level neuroanatomical changes that provide unique insights into disease progression and biological vulnerability.

MIND-based structural similarity quantifies how morphometrically alike a given brain region is to all others, integrating multiple features of cortical architecture. A region’s MIND degree thus reflects its structural integration within the cortex. Normative modelling of MIND degree allowed us to assess whether regional structural similarity was higher or lower than expected based on typical patterns in healthy adults.

Our main findings show that negative deviations were highly sensitive to different aspects of Alzheimer’s disease pathology, effectively stratifying clinical stages. The amount of negative deviations increased progressively—from individuals with stable mild cognitive impairment, to those with progressive cognitive decline, and lastly to those with a diagnosis of Alzheimer’s disease. Our APOE ε4 analyses also revealed strong sex-specific effects, with ε4 homozygote females showing substantially more negative deviations than other genotype groups. Spatially, deviations co-localised with neurobiological properties, particularly neurotransmitter receptor density maps and some axes of cortical organization. Crucially, in terms of prognosis, although there was no significant effect between deviations and mortality, individuals with more negative deviations showed greater postmortem neuropathological burden, establishing a direct link between our in-vivo neuroimaging markers and underlying disease pathology.

Importantly, our overlap analyses revealed low regional concordance across individual patients (<10%), indicating substantial individual heterogeneity that traditional group-average approaches would overlook. This heterogeneity suggests that clinical applications may benefit from individualised deviation approaches, while highlighting the value of population modelling for detecting meaningful patterns amid this individual variability.

### 4.2 Disrupted Cortical Integration in Alzheimer’s Disease

Our findings contribute to a growing body of work suggesting that Alzheimer’s disease may reflect not only regional atrophy, but also disruptions in the coordinated structural organisation of the cortex^48,49^. The predominance of negative deviations points to a possible breakdown in inter-regional structural relationships. Notably, significant negative deviations were predominantly located in association cortical areas along the sensorimotor-association cortical axis^41^, while positive ones more in primary sensory regions. Our approach extends beyond conventional connectivity measures by capturing multivariate morphometric similarity, which may reflect deeper principles of cortical maturation and architectural coordination^4^. Our findings may reflect a disruption of homophilic cortical architecture, i.e., the principle that structurally and transcriptionally similar regions are preferentially aligned and coordinated in the brain^4^. While further validation is needed, these results support the view that structural disintegration at the network level may be a relevant dimension of Alzheimer’s pathophysiology.

The network-level enrichment of negative deviations, particularly in the default mode, attention, and frontoparietal networks, provides support for the network degeneration hypothesis^50^. However, our findings also demonstrate selective vulnerability patterns—while somatomotor networks showed significant negative deviation enrichment, the visual network remained spared. The contrast between affected somatomotor networks and spared visual networks suggests that MIND-based measures may detect early disruption of structural alignment in systems that are otherwise considered more functionally resilient in dementia. Importantly, while our regional overlap analyses showed low rates across individual patients, network-level analyses revealed that when regions were grouped into functional systems, preferential targeting patterns emerged. This suggests that AD may affect specific networks consistently, but with individual variation in which particular regions within those networks show deviations. This pattern of network-level consistency amid regional and inter-individual heterogeneity would be missed by traditional approaches that focus on group-average regional effects.

Spatial co-location analyses between structural deviations and different neuroanatomical maps provided biological context for the observed results. In AD, regions with *decreased* morphometric similarity overlapped with areas of high serotonergic (5-HT2a, 5-HT4), GABAergic, and glutamatergic (mGluR5) receptor density. Although these represent group-averaged receptor maps, these findings are consistent with emerging evidence for neurotransmitter system dysfunction in AD^51,52^ and suggest that MIND-based deviations may be sensitive to disruption in regions supporting modulatory network coordination^36^. Interestingly, in mild cognitive impairment (MCI)-Stable participants, we observed the opposite pattern: regions with higher-than-expected structural similarity aligned with areas rich in mGluR5, MOR, and GABAergic receptors (alongside many others, spanning from nicotinic acetylcholine receptor to dopaminergic ones and more). Taken together, this pattern—positive deviations in MCI stages, shifting to negative deviations in AD—may reflect a transition from structural resilience to breakdown as the disease progresses, although no direct causal link can be made here. The observed results suggest that structural deviations can be biologically decoded by taking into account underlying neurotransmitter systems^36,43^.

We also observed stage-specific relationships with other cortical organisational properties. In MCI, regions exhibiting positive deviations corresponded with areas of high evolutionary expansion^53^ and positive allometric scaling^54^, which are regions typically associated with cognitive flexibility and prolonged neurodevelopment^55^. Conversely, in AD, these associations reversed, with negative deviations predominantly affecting these same regions, potentially suggesting that evolutionarily expanded and late-maturing areas may become selectively vulnerable as the disease progresses. This pattern is consistent with the “last-in, first-out” principle^56^, whereby brain regions that develop relatively late during adolescence also show accelerated degeneration in aging and heightened vulnerability to neurodegenerative diseases^56^. Our findings suggest that normative modelling of MIND networks may capture these vulnerability patterns at the individual level, as regions transition from showing resilience in MCI to exhibiting pathology in AD.

### 4.3 Genetic Architecture and Sex-Specific Vulnerability

Our APOE findings reveal a nuanced picture of genetic risk and disease-related brain changes, with a marked sex-specificity of APOE ε4 effects on negative deviations. Our results suggest sex-specific vulnerability, particularly among ε4 homozygous females, in the form of increased number of negative deviations. This finding aligns with evidence that APOE ε4 risk for AD is greater in women than men^57,58^. This could not be explained by group size differences or imbalances, suggesting fundamental differences in how genetic risk manifests structurally. Our findings provide some preliminary evidence that this vulnerability manifests as measurable disruption of morphometric similarity networks. Interestingly, ε2 carriers showed significantly elevated positive deviations compared to ε3 homozygotes, potentially representing some neuroimaging evidence of ε2’s protective effects at the network level. While the underlying mechanisms remain to be clarified, this finding is consistent with evidence for ε2’s protective role in AD^59^.

### 4.4 Prognostic Implications and Clinical Translation

In the context of clinical application, the observed association between structural deviation burden and postmortem neuropathological severity suggests that morphometric similarity networks may hold promise as biologically informed markers of disease burden. These measures could contribute to more refined patient stratification in research settings, support efforts to stage disease progression, and inform individualised assessments. The prognostic value of our normative modelling-derived measures is consistent with recent longitudinal work showing that increasing outlier burden over time predicts clinical progression to dementia^18^. However, further validation is required to determine appropriate thresholds for clinical use, assess reproducibility across diverse cohorts and imaging protocols, and establish how these measures compare to existing imaging biomarkers. While we also present group-level analyses to validate our approach, the core strength of our framework lies in calculating individual-level deviation maps. This directly addresses AD heterogeneity and enables precision medicine approaches for personalised patient assessment and prognosis.

### 4.5 Limitations and Future Work

Several limitations should be acknowledged. First, while our cross-sectional approach reveals individual-level deviations at single timepoints, it prevents direct assessment of temporal changes. Longitudinal studies will be necessary to clarify how MIND-based deviations evolve over time and establish their prognostic value, as shown for traditional morphometric measures^15^. Second, the choice of morphometric features inevitably influenced network construction, and one should note that using alternative features might provide different biological insights. Although we selected our features based on those widely used in previous work^5,7,60^, it remains an open question how many, and which exact number and features is the “optimal” choice^4^. Additionally, MIND networks were restricted to cortical regions, and it would be useful to extend analyses to the subcortex^61^. Third, our cohort consisted primarily of individuals of European ancestry, potentially limiting generalisability to other populations and requiring validation across diverse groups before broad clinical application. Related to this, validating our combined normative MIND approach on portable low-field MRI scanners^62^ will be important to ensure its accessibility and scalability in real-world clinical settings, supporting broader adoption across diverse healthcare environments. Finally, while our correlations with neuropathological measures and spatial co-location analyses are encouraging, more direct validation through post-mortem studies or molecular imaging would strengthen mechanistic interpretations.

### 4.6 Conclusion

This study introduces a novel hybrid framework that integrates MIND networks with hierarchical Bayesian normative modelling to map individual-level deviations in cortical structure. By moving beyond traditional univariate analyses, our multivariate approach captures biologically grounded markers of individual brain disruption in Alzheimer’s disease. This framework not only stratifies disease stages and genetic risk but also links neuroanatomical variation to severity of postmortem pathology. It also allowed us to spatially decode the results with maps of neurotransmitter receptor density and specific cortical organization properties. Taken together, these findings highlight the promise of multivariate, network-based normative models as scalable tools for precision mapping of brain disorders.

## Data Availability

Data from the National Alzheimer's Coordinating Center (NACC) are publicly available and can be requested through the NACC website (https://naccdata.org/requesting-data/submit-data-request).
UK Biobank Data were obtained from UK Biobank (www.ukbiobank.ac.uk). Access to UK Biobank data can be requested from the UK Biobank Access Management Team.

## Acknowledgements

This research has been conducted using the UK Biobank Resource under Application Number 20904. Data were curated and analysed using a computational facility funded by an MRC research infrastructure award (MR/M009041/1) to the School of Clinical Medicine, University of Cambridge and supported by the mental health theme of the NIHR Cambridge Biomedical Research Centre. The views expressed are those of the authors and not necessarily those of the NIH, NHS, the NIHR or the Department of Health and Social Care.

The NACC database is funded by NIA/NIH Grant U24 AG072122. NACC data are contributed by the NIA-funded ADRCs: P30 AG062429 (PI James Brewer, MD, PhD), P30 AG066468 (PI Oscar Lopez, MD), P30 AG062421 (PI Bradley Hyman, MD, PhD), P30 AG066509 (PI Thomas Grabowski, MD), P30 AG066514 (PI Mary Sano, PhD), P30 AG066530 (PI Helena Chui, MD), P30 AG066507 (PI Marilyn Albert, PhD), P30 AG066444 (PI David Holtzman, MD), P30 AG066518 (PI Lisa Silbert, MD, MCR), P30 AG066512 (PI Thomas Wisniewski, MD), P30 AG066462 (PI Scott Small, MD), P30 AG072979 (PI David Wolk, MD), P30 AG072972 (PI Charles DeCarli, MD), P30 AG072976 (PI Andrew Saykin, PsyD), P30 AG072975 (PI Julie A. Schneider, MD, MS), P30 AG072978 (PI Ann McKee, MD), P30 AG072977 (PI Robert Vassar, PhD), P30 AG066519 (PI Frank LaFerla, PhD), P30 AG062677 (PI Ronald Petersen, MD, PhD), P30 AG079280 (PI Jessica Langbaum, PhD), P30 AG062422 (PI Gil Rabinovici, MD), P30 AG066511 (PI Allan Levey, MD, PhD), P30 AG072946 (PI Linda Van Eldik, PhD), P30 AG062715 (PI Sanjay Asthana, MD, FRCP), P30 AG072973 (PI Russell Swerdlow, MD), P30 AG066506 (PI Glenn Smith, PhD, ABPP), P30 AG066508 (PI Stephen Strittmatter, MD, PhD), P30 AG066515 (PI Victor Henderson, MD, MS), P30 AG072947 (PI Suzanne Craft, PhD), P30 AG072931 (PI Henry Paulson, MD, PhD), P30 AG066546 (PI Sudha Seshadri, MD), P30 AG086401 (PI Erik Roberson, MD, PhD), P30 AG086404 (PI Gary Rosenberg, MD), P20 AG068082 (PI Angela Jefferson, PhD), P30 AG072958 (PI Heather Whitson, MD), P30 AG072959 (PI James Leverenz, MD)

## Consent Statement

All data collection protocols were approved by the institutional review board of each cohort, and informed written consent was obtained from all participants in the original studies.

