## Supplementary Material for "Structural Similarity Networks Reveal Brain Vulnerability in Dementia"

### 1. **Normative Modelling Implementation and Quality Controls**

**Model Configuration and Specifications**

We implemented a hierarchical Bayesian regression model using the PCN toolkit (<https://github.com/amarquand/PCNtoolkit> ) with a specific configuration to optimize performance and balance model complexity. The model used *bspline* basis functions with linear terms for both mean (μ) and variance (σ) parameters. We included random intercepts for the mean to appropriately account for site effects, while excluding random slopes and sigma random effects to maintain model parsimony and stability. For sampling, we utilized the No-U-Turn Sampler (NUTS) with these specifications: Number of chains: 4; Samples per chain: 5000; Tuning samples: 2000; Target acceptance rate: 0.99. We trained normative models on Morphometric INverse Divergence (MIND) weighted degree values from 35,133 UK Biobank participants to serve as our reference dataset. For the NACC dataset, we assigned healthy controls to both the transfer/adaptation set (HCtrain; N=1624) and the test set (HCtest; N=406) sets in an 80:20 ratio. The HCtrain set was used for model recalibration, while the HCtest set along with patient groups (AD: N=624; MCI Stable: N=643; MCI Progressive: N=300) were used for assessing deviations from normative brain structure.

**MCMC Convergence Assessment**
To assess the convergence of our HBR normative models, the Gelman–Rubin statistic ($\hat{R}$) was computed for all model parameters in line with what recommended for the PCNToolkit (see available tutorial online at the following link: <https://github.com/predictive-clinical-neuroscience/PCNtoolkit-demo/blob/81967c1e158feeaa3651aed6decee80f3018a563/tutorials/HBR_SHASH/HBR_Tutorial.ipynb>). $\hat{R}$ quantifies agreement between chains by comparing within-chain and between-chain variance. Ideal convergence corresponds to an $\hat{R}$ value of 1.00, while values below 1.05, or a more stringent 1.01 used here, are widely accepted as evidence of good convergence (Vehtari et al., 2021). As a representative example, we visualized $\hat{R}$ values across all parameter groups for one model (see Supplementary Figure 1). Most parameters across mean and variance components had $\hat{R}$ values well below 1.01, indicating good mixing. A small number of early spline coefficients (e.g., in sigma_intercept_mu and offset_slope_mu) slightly exceeded 1.01 but remained below 1.05, likely reflecting minor local uncertainties rather than substantive convergence failures. No parameters showed evidence of non-convergence ($\hat{R}$ > 1.1). These findings confirm the overall stability and reliability of the posterior estimates used in downstream analyses.

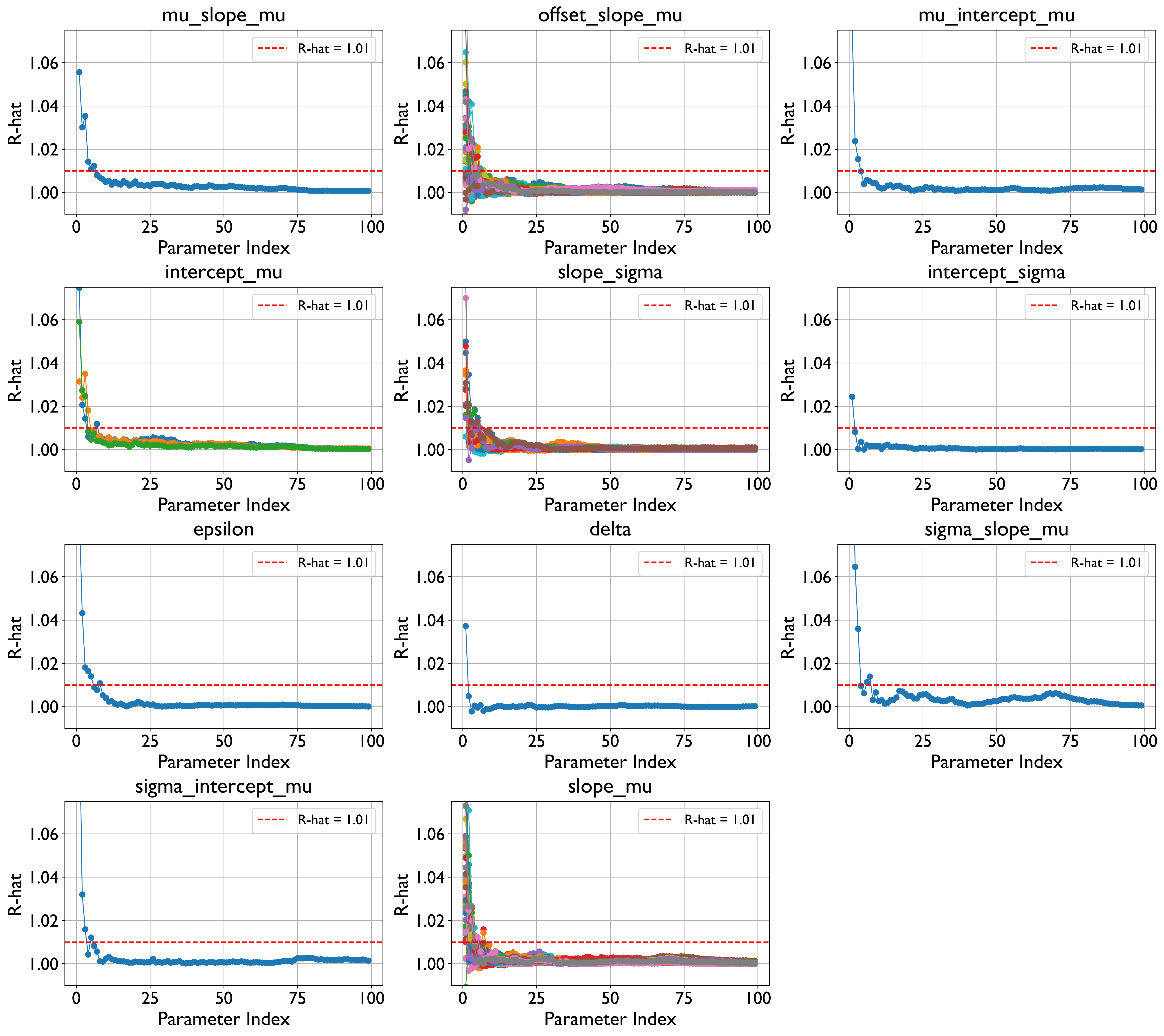

**Supplementary Figure 1-** $\hat{R}$ convergence plots for all model parameters from one representative ROI model. The red line indicates the $\hat{R}$= 1.01 threshold. Most parameters show values at or near 1.00.

**Data Preparation and Site Handling**

To facilitate proper handling of multi-site data, unique site identifiers were assigned to each location, creating batch effect matrices for both the training and test sets. This approach allowed the model to parse out site variability effectively while maintaining the ability to detect true biological differences. All sites included in the test set were also represented in the training set to ensure proper calibration of site-specific parameters.

**Model Performance Evaluation**

We evaluated the explained variance of the model to assess its performance across regions. Variance plots were generated to verify the distribution of Explained Variance (which quantifies the proportion of variance in the ROI that is explained by the model) and Standardized Mean Squared Error (measures the average squared difference between the predicted and observed values, standardized by the variance in the data). See Supplementary Figure 2.

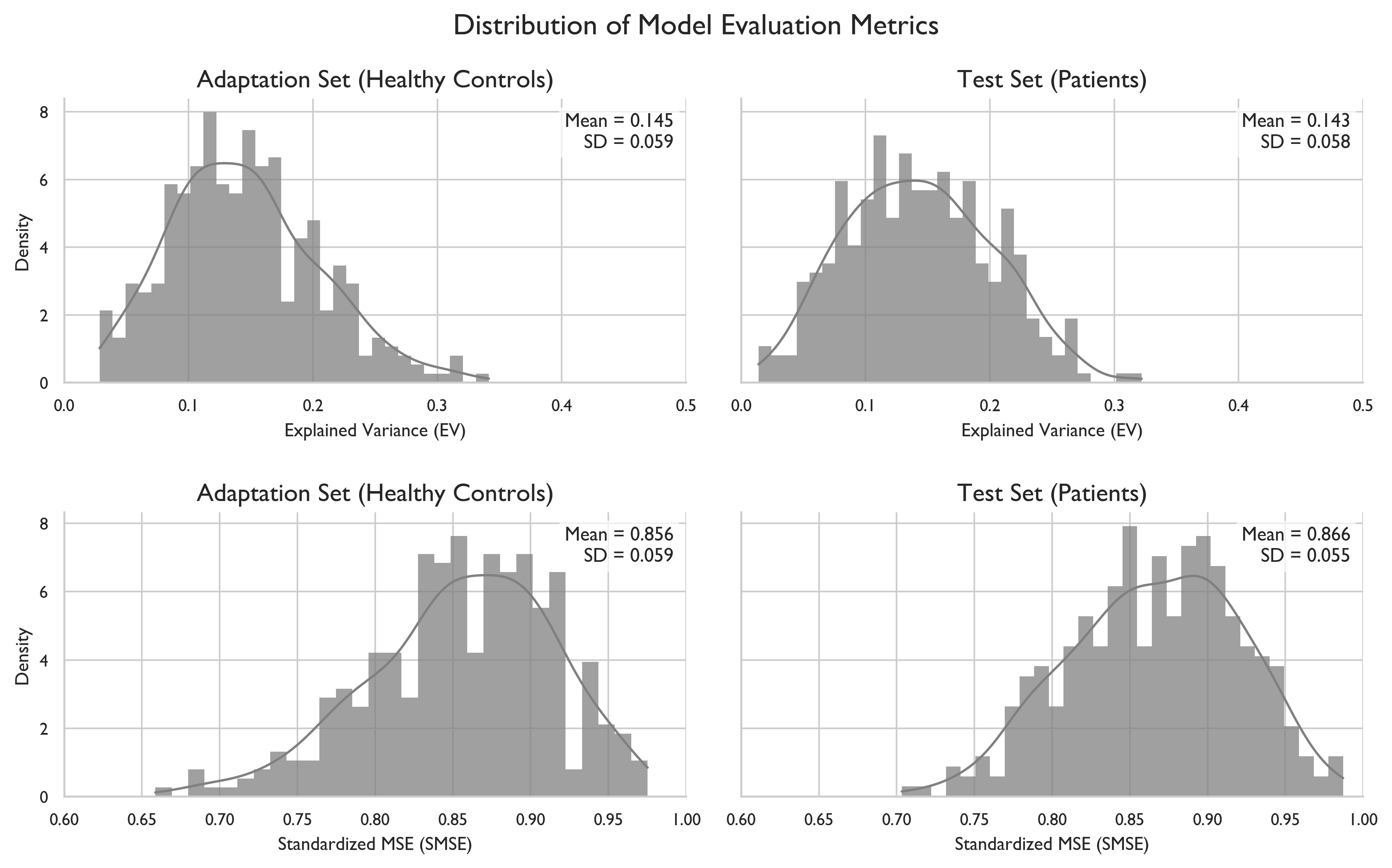

**Supplementary Figure 2**- Distribution of Model Evaluation The distributions of i) explained variance (EXPV; higher is better) in the top two rows, and ii) Standardized mean squared error (SMSE; lower is better) in the lower panel, across 360 ROIs in the trasfer/adaptation sample and in the test cohort.

We generated age distribution density plots for controls in the UK Biobank training set and both the adaptation/transfer set (HC_train_) and test set of the NACC dataset (AD, MCI Stable, MCI Progressive, HC_test_) to ensure appropriate age coverage across cohorts. See panels in Supplementary Figure 3 below. We also plotted age distributions by site and gender for both sets to ensure no demographic or site-related imbalances. In addition, site-wise distributions of participants’ age in the adaptation and test samples were plotted for each site. See Supplementary Figure 4.

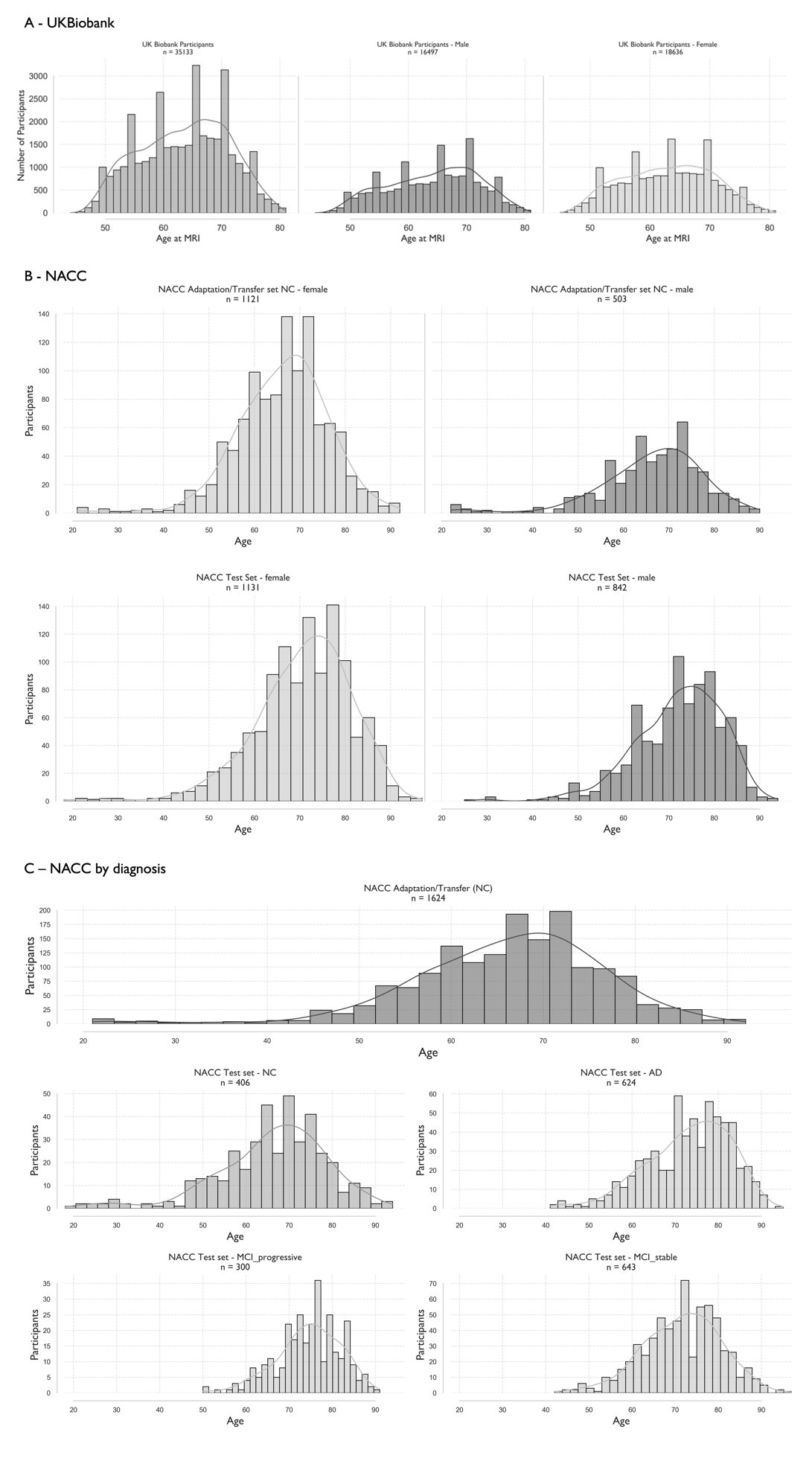

**Supplementary Figure 3** – (**A)** Histograms showing age distributions of participants from the UK Biobank normative model training set, stratified by sex. **(B)** Age distributions for the NACC adaptation (transfer) and test cohorts, stratified by sex. Kernel density estimates (KDEs) are overlaid to visualize distribution shape. **(C)** Age distributions in the NACC test set stratified by diagnostic group: Alzheimer's Disease (AD), Mild Cognitive Impairment (MCI; stable and progressive), and cognitively normal controls. These plots confirm appropriate age coverage and diagnostic representation in the model adaptation and test sets.

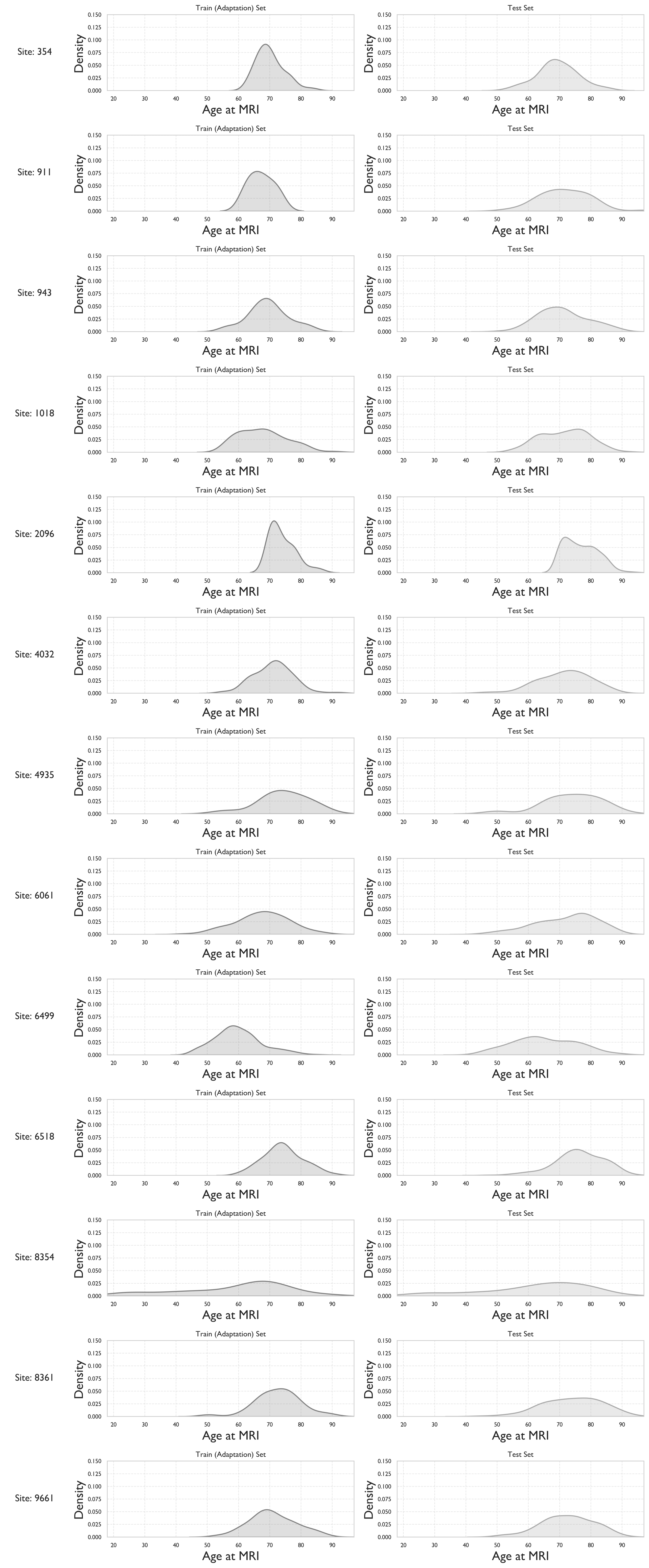

**Supplementary Figure 4**- Site-wise distributions of participant age at MRI scan in the adaptation and test NACC cohort. For each site, the kernel density estimate (KDE) of age is shown separately for the adaptation set (on the left) and test set (on the right).

**Controlling for Total Intracranial Volume in Normative Modelling**

Initial analysis revealed significant correlations between estimated Total Intracranial Volume (eTIV) and the number of outlier regions (defined as z-scores exceeding ±1.96) across diagnostic groups. These correlations could potentially confound interpretation of disease-related abnormalities since variations might be partly attributable to head size rather than pathology. See Supplementary Figure 5 below.

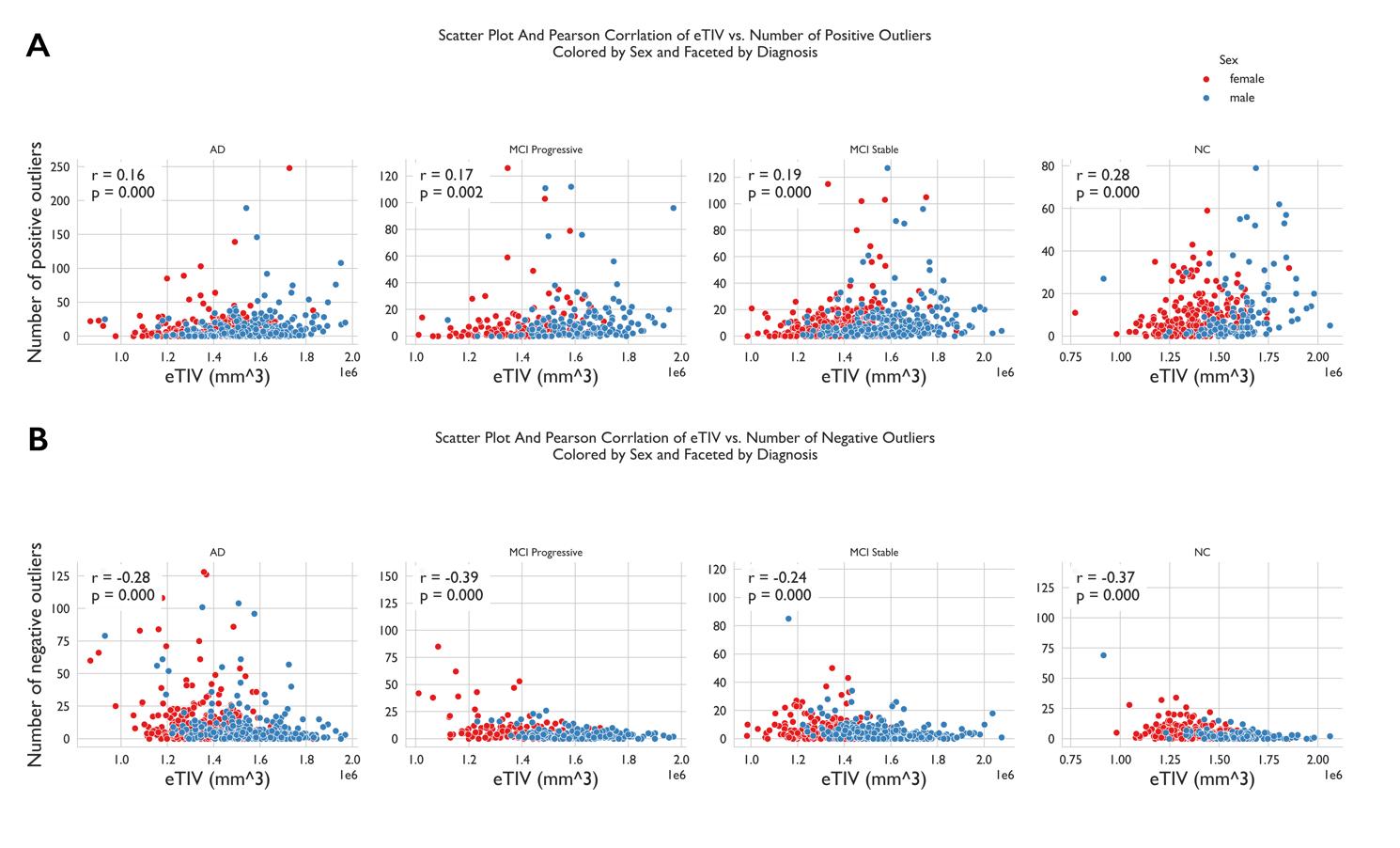

**Supplementary Figure 5** - **Correlation between estimated total intracranial volume (eTIV) and cortical deviation burden across diagnostic groups.** (A) Scatter plots showing the relationship between eTIV and the number of extreme positive deviations (Z > 1.96) for each diagnostic group: Alzheimer's Disease (AD), MCI Progressive, MCI Stable, and cognitively normal controls (NC). Points are coloured by sex, and Pearson correlation coefficients are reported per group. Significant positive correlations were observed in AD and MCI Progressive groups, suggesting that head size may confound deviation estimates if uncorrected. (B) Equivalent plots for negative deviations (Z < –1.96). Correlation strengths and directions vary by group and sex, reinforcing the necessity of residualizing regional deviation scores for eTIV to mitigate bias.

We first ensured removal of outliers in eTIV for each diagnosis and sex (defined as being outside the IQR range). This led to the removal of 19 females (AD: 5, MCI Stable: 9, NC: 5) and N=11 males (AD: 1, MCI Progressive: 1, MCI Stable: 2, NC: 7). We subsequently performed residualisation separately for each demographic group (defined again by sex and diagnosis) to account for potentially different relationships between brain measures and head size across these populations. For each brain region and demographic group, we fit a linear regression model predicting the regional z-score from eTIV, calculated residuals (observed z-score minus predicted z-score), added back the group mean to maintain the original scale and interpretability, and verified the effectiveness of residualisation by testing for remaining correlations. See Supplementary Figure 6 below.

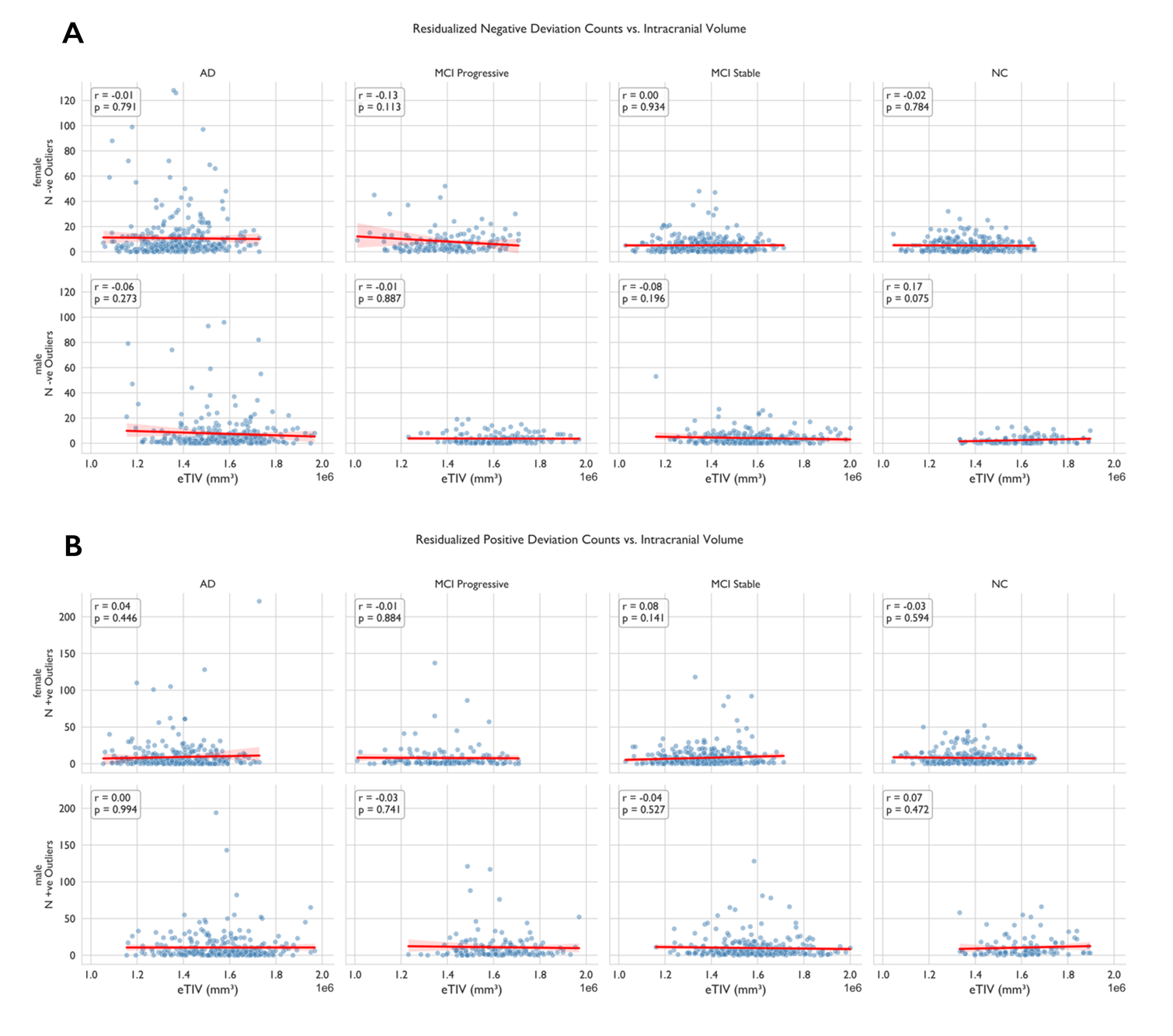

**Supplementary Figure 6**- Relationship between residualised deviation counts and intracranial volume after eTIV correction. (A) Scatter plots showing the association between eTIV and residualised negative deviation counts (Z < –1.96) across diagnostic groups following group- and sex-specific eTIV adjustment. Each panel displays Pearson’s correlation coefficient (r) and corresponding p-value. No significant associations were observed, confirming effective removal of eTIV-related confounding. (B) Equivalent analysis for residualised positive deviation counts.

This group-specific approach was chosen over global residualisation because relationships between brain structure and head size often differ between males and females. After residualisation, the correlation between eTIV and deviation counts was substantially reduced (average correlation coefficient reduction: 0.24 for positive deviations, 0.27 for negative deviations), with most correlations no longer statistically significant. The residualised z-scores provide a more specific representation of disease-related brain abnormalities, adjusting for the confounding effect of head size while preserving the interpretability of the normative modelling framework. All subsequent analyses used these residualised values to ensure that reported group differences reflect pathological processes rather than anatomical variations associated with head size.

**Evaluation of Age-Related Bias in Deviation Estimates**

Lastly, we assessed the correlation between age and deviation burden, quantified as the mean absolute Z-score across all regions of interest, in the final test set. This was done to evaluate whether model predictions were biased by age, particularly at the extremes of the age distribution, where model uncertainty tends to be greater due to fewer observations. Results indicated no significant correlation between age and deviations (Pearson’s *r* = -0.026, *p* = 0.250), suggesting that individual-level deviation estimates were not systematically influenced by age-related factors.

### 2. **Relationship Between MIND Networks and Morphometric Features**

To investigate the anatomical correlates of MIND-derived deviation maps, we computed regional Pearson correlations between weighted degree values and the five morphometric MRI features derived from FreeSurfer that were used to construct the MIND networks, namely: cortical thickness (CT), mean curvature (MC), sulcal depth (SD), surface area (SA), and grey matter volume (GMV), across 360 Glasser regions and separately for each diagnostic group (Healthy Controls (NC), MCI Stable, MCI Progressive, and AD). These features were chosen in line with previous work. As shown in Supplementary Figure 7 below, the strongest and most consistent associations were observed with GMV (r ≈ 0.45) and SA (r ≈ 0.38), followed by robust negative correlations with SD (r ≈ –0.39). These effects were statistically significant across all groups, with FDR-corrected p-values. Correlations with MC (r ≈ 0.16–0.20) were weaker but remained significant. The only non-significant feature was CT, potentially because it did not provide further information in addition to SA and regional volume. These findings support the anatomical interpretability of MIND networks.

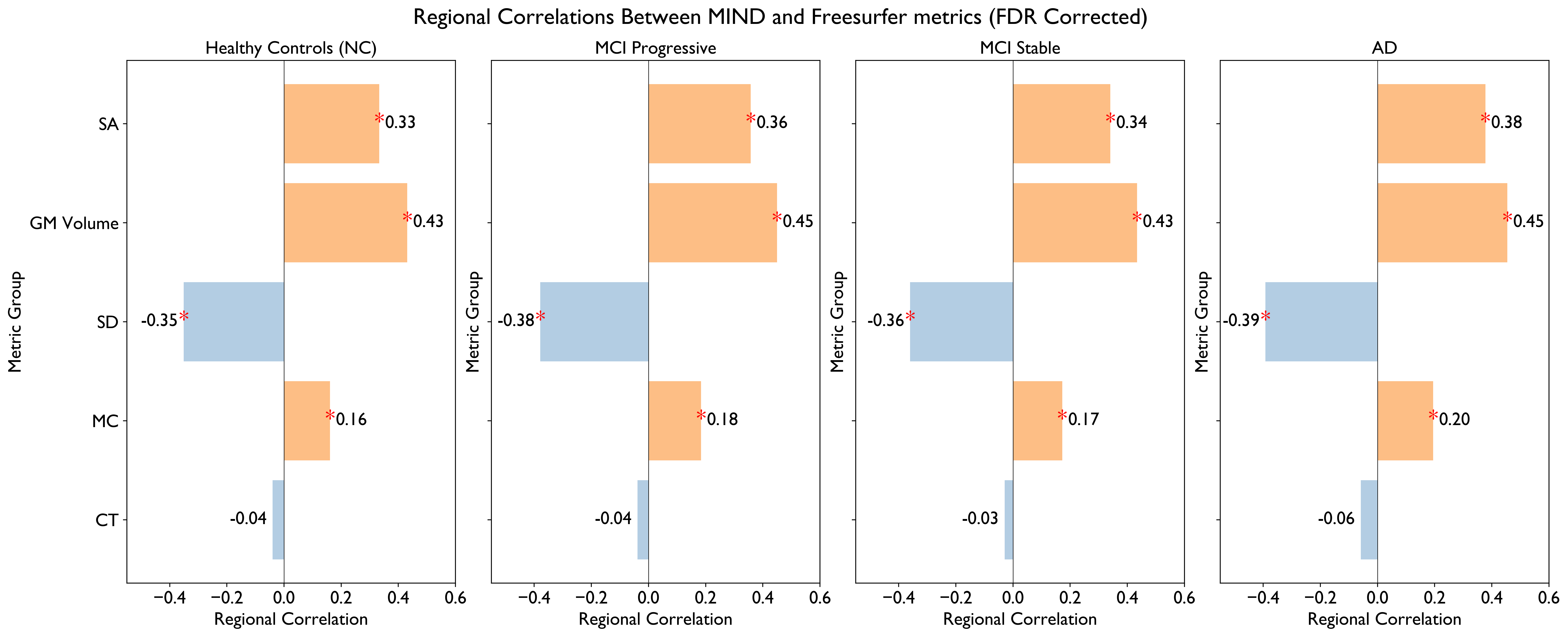

**Supplementary Figure 7** – Pearson Correlations Between MIND and morphometric MRI Metrics derived from Freesurfer and used to construct the MIND networks. Asterisks (*) indicate significant correlations (P-val after FDR correction).

### 3. **Group ANOVA For Positive and Negative Deviation Counts**

Supplementary Table 1 provides detailed statistical outputs for group comparisons of deviation counts. One-way ANOVAs were performed separately for positive and negative deviations, followed by Tukey’s HSD post-hoc tests to assess pairwise group differences. Positive and negative deviation counts were calculated as the number of cortical regions with Z-scores greater than 1.96 or less than -1.96, respectively. The analysis included all cortical regions assessed using the MIND normative model. Full results, including confidence intervals and adjusted p-values, are shown in Supplementary Tables 1 and 2 below.

**Supplementary Table 1**- ANOVA and Tukey HSD post-hoc test results for positive and negative deviation counts across diagnostic groups.

| **Deviation Type** | **Comparison** | **Mean Diff** | **Lower CI** | **Upper CI** | **p-adj** | **Significant** | **ANOVA F** | **ANOVA p-value** |
| --- | --- | --- | --- | --- | --- | --- | --- | --- |
| **Positive** | AD vs. MCI Progressive | -0.42 | -3.36 | 2.53 | 0.98 | No | 0.59 | 0.62 |
|  | AD vs. MCI Stable | -1.11 | -3.47 | 1.26 | 0.62 | No |  |  |
|  | AD vs. NC | -1.03 | -3.73 | 1.66 | 0.76 | No |  |  |
|  | MCI Progressive vs. MCI Stable | -0.69 | -3.62 | 2.24 | 0.93 | No |  |  |
|  | MCI Progressive vs. NC | -0.62 | -3.82 | 2.59 | 0.96 | No |  |  |
|  | MCI Stable vs. NC | 0.07 | -2.61 | 2.75 | 1.00 | No |  |  |
| **Negative** | AD vs. MCI Progressive | -3.04 | -5.13 | -0.95 | 0.00 | Yes | 24.39 | <0.001 |
|  | AD vs. MCI Stable | -4.83 | -6.51 | -3.14 | <0.001 | Yes |  |  |
|  | AD vs. NC | -5.33 | -7.24 | -3.41 | <0.001 | Yes |  |  |
|  | MCI Progressive vs. MCI Stable | -1.78 | -3.87 | 0.30 | 0.12 | No |  |  |
|  | MCI Progressive vs. NC | -2.28 | -4.56 | 0.00 | 0.05 | Yes |  |  |
|  | MCI Stable vs. NC | -0.50 | -2.41 | 1.41 | 0.91 | No |  |  |

**Supplementary Table 2 –** Full details and p-values of the Cohen’s d analyses with FDR correction.

| **ROI  (lh, rh  hemi)** | **Comparison** | **Less  Severe Group** | **More  Severe  Group** | **Mean  Less  Severe** | **Mean  More  Severe** | **Cohens d observed** | **P FDR** |
| --- | --- | --- | --- | --- | --- | --- | --- |
| lhV1 | MCI Progressive_vs_AD | MCI Progressive | AD | -0.099 | 0.261 | -0.335 | 0.0090 |
| rhV1 | MCI Progressive_vs_AD | MCI Progressive | AD | -0.140 | 0.206 | -0.324 | 0.0090 |
| rh3b | MCI Progressive_vs_AD | MCI Progressive | AD | 0.116 | 0.427 | -0.309 | 0.0090 |
| lh3b | MCI Progressive_vs_AD | MCI Progressive | AD | 0.155 | 0.436 | -0.277 | 0.0120 |
| lh23c | MCI Progressive_vs_AD | MCI Progressive | AD | 0.224 | -0.027 | 0.241 | 0.0432 |
| rhd32 | MCI Progressive_vs_AD | MCI Progressive | AD | 0.279 | 0.017 | 0.248 | 0.0320 |
| lh24dv | MCI Progressive_vs_AD | MCI Progressive | AD | 0.151 | -0.094 | 0.249 | 0.0315 |
| lh47s | MCI Progressive_vs_AD | MCI Progressive | AD | 0.233 | -0.030 | 0.258 | 0.0309 |
| rhFOP2 | MCI Progressive_vs_AD | MCI Progressive | AD | 0.176 | -0.086 | 0.262 | 0.0120 |
| lhFOP3 | MCI Progressive_vs_AD | MCI Progressive | AD | 0.222 | -0.075 | 0.303 | 0.0090 |
| lh24dv | MCI Stable_vs_AD | MCI Stable | AD | 0.317 | -0.094 | 0.434 | 0.0006 |
| lhp32pr | MCI Stable_vs_AD | MCI Stable | AD | 0.295 | -0.073 | 0.387 | 0.0006 |
| lhFOP3 | MCI Stable_vs_AD | MCI Stable | AD | 0.281 | -0.075 | 0.366 | 0.0006 |
| lh23c | MCI Stable_vs_AD | MCI Stable | AD | 0.337 | -0.027 | 0.362 | 0.0006 |
| rhFOP2 | MCI Stable_vs_AD | MCI Stable | AD | 0.264 | -0.086 | 0.361 | 0.0006 |
| rha32pr | MCI Stable_vs_AD | MCI Stable | AD | 0.362 | 0.030 | 0.346 | 0.0006 |
| rhOP2-3 | MCI Stable_vs_AD | MCI Stable | AD | 0.210 | -0.117 | 0.338 | 0.0006 |
| lhFOP2 | MCI Stable_vs_AD | MCI Stable | AD | 0.277 | -0.033 | 0.321 | 0.0006 |
| rh24dv | MCI Stable_vs_AD | MCI Stable | AD | 0.226 | -0.085 | 0.317 | 0.0006 |
| lhp24 | MCI Stable_vs_AD | MCI Stable | AD | 0.360 | 0.059 | 0.316 | 0.0006 |
| lhPHA2 | MCI Stable_vs_AD | MCI Stable | AD | 0.244 | -0.059 | 0.314 | 0.0006 |
| rhp32 | MCI Stable_vs_AD | MCI Stable | AD | 0.280 | -0.018 | 0.311 | 0.0006 |
| lh31pd | MCI Stable_vs_AD | MCI Stable | AD | 0.239 | -0.047 | 0.307 | 0.0006 |
| rhPOS2 | MCI Stable_vs_AD | MCI Stable | AD | 0.265 | -0.033 | 0.302 | 0.0006 |
| rhd32 | MCI Stable_vs_AD | MCI Stable | AD | 0.322 | 0.017 | 0.299 | 0.0006 |
| lhPOS2 | MCI Stable_vs_AD | MCI Stable | AD | 0.235 | -0.054 | 0.298 | 0.0006 |
| rhPGi | MCI Stable_vs_AD | MCI Stable | AD | 0.204 | -0.067 | 0.294 | 0.0006 |
| rhFOP3 | MCI Stable_vs_AD | MCI Stable | AD | 0.222 | -0.055 | 0.289 | 0.0006 |
| lha24 | MCI Stable_vs_AD | MCI Stable | AD | 0.314 | 0.035 | 0.285 | 0.0006 |
| lhd32 | MCI Stable_vs_AD | MCI Stable | AD | 0.179 | -0.103 | 0.283 | 0.0006 |
| lha24pr | MCI Stable_vs_AD | MCI Stable | AD | 0.301 | 0.040 | 0.279 | 0.0010 |
| rhIP2 | MCI Stable_vs_AD | MCI Stable | AD | 0.119 | -0.147 | 0.275 | 0.0006 |
| lhFOP4 | MCI Stable_vs_AD | MCI Stable | AD | 0.241 | -0.021 | 0.273 | 0.0006 |
| rhIP0 | MCI Stable_vs_AD | MCI Stable | AD | 0.150 | -0.107 | 0.271 | 0.0006 |
| lhp24pr | MCI Stable_vs_AD | MCI Stable | AD | 0.259 | 0.003 | 0.271 | 0.0006 |
| rhPHA2 | MCI Stable_vs_AD | MCI Stable | AD | 0.267 | 0.011 | 0.271 | 0.0006 |
| rh31pd | MCI Stable_vs_AD | MCI Stable | AD | 0.183 | -0.075 | 0.271 | 0.0006 |
| lh6a | MCI Stable_vs_AD | MCI Stable | AD | 0.202 | -0.048 | 0.267 | 0.0006 |
| rhp32pr | MCI Stable_vs_AD | MCI Stable | AD | 0.247 | -0.011 | 0.267 | 0.0006 |
| rha24 | MCI Stable_vs_AD | MCI Stable | AD | 0.373 | 0.108 | 0.265 | 0.0006 |
| rh23c | MCI Stable_vs_AD | MCI Stable | AD | 0.309 | 0.041 | 0.264 | 0.0006 |
| rhAVI | MCI Stable_vs_AD | MCI Stable | AD | 0.154 | -0.098 | 0.262 | 0.0006 |
| lh24dd | MCI Stable_vs_AD | MCI Stable | AD | 0.173 | -0.078 | 0.258 | 0.0006 |
| lh47s | MCI Stable_vs_AD | MCI Stable | AD | 0.223 | -0.030 | 0.257 | 0.0006 |
| lh31a | MCI Stable_vs_AD | MCI Stable | AD | 0.255 | 0.008 | 0.254 | 0.0006 |
| rh24dd | MCI Stable_vs_AD | MCI Stable | AD | 0.230 | -0.011 | 0.249 | 0.0006 |
| rhp24pr | MCI Stable_vs_AD | MCI Stable | AD | 0.270 | 0.027 | 0.248 | 0.0006 |
| lhAVI | MCI Stable_vs_AD | MCI Stable | AD | 0.120 | -0.128 | 0.248 | 0.0006 |
| lhPFm | MCI Stable_vs_AD | MCI Stable | AD | 0.128 | -0.123 | 0.248 | 0.0006 |
| lhPI | MCI Stable_vs_AD | MCI Stable | AD | 0.239 | 0.004 | 0.246 | 0.0006 |
| lha32pr | MCI Stable_vs_AD | MCI Stable | AD | 0.186 | -0.058 | 0.242 | 0.0006 |
| rh9p | MCI Stable_vs_AD | MCI Stable | AD | 0.185 | -0.049 | 0.241 | 0.0006 |
| lhV3CD | MCI Stable_vs_AD | MCI Stable | AD | 0.135 | -0.104 | 0.241 | 0.0006 |
| rhPGs | MCI Stable_vs_AD | MCI Stable | AD | 0.126 | -0.117 | 0.241 | 0.0006 |
| rhPFm | MCI Stable_vs_AD | MCI Stable | AD | 0.189 | -0.043 | 0.239 | 0.0006 |
| lhp32 | MCI Stable_vs_AD | MCI Stable | AD | 0.214 | -0.017 | 0.239 | 0.0006 |
| rhSCEF | MCI Stable_vs_AD | MCI Stable | AD | 0.106 | -0.118 | 0.235 | 0.0010 |
| lhIP0 | MCI Stable_vs_AD | MCI Stable | AD | 0.108 | -0.119 | 0.234 | 0.0006 |
| lhTPOJ2 | MCI Stable_vs_AD | MCI Stable | AD | 0.177 | -0.050 | 0.229 | 0.0006 |
| rhTPOJ2 | MCI Stable_vs_AD | MCI Stable | AD | 0.217 | -0.002 | 0.228 | 0.0010 |
| rh47l | MCI Stable_vs_AD | MCI Stable | AD | 0.224 | 0.018 | 0.224 | 0.0010 |
| rh31a | MCI Stable_vs_AD | MCI Stable | AD | 0.253 | 0.031 | 0.223 | 0.0013 |
| rh9m | MCI Stable_vs_AD | MCI Stable | AD | 0.187 | -0.030 | 0.221 | 0.0017 |
| rhIPS1 | MCI Stable_vs_AD | MCI Stable | AD | 0.200 | -0.016 | 0.221 | 0.0006 |
| rh10v | MCI Stable_vs_AD | MCI Stable | AD | 0.239 | 0.026 | 0.219 | 0.0013 |
| rhPI | MCI Stable_vs_AD | MCI Stable | AD | 0.250 | 0.057 | 0.215 | 0.0010 |
| rh25 | MCI Stable_vs_AD | MCI Stable | AD | 0.219 | 0.004 | 0.214 | 0.0010 |
| lh52 | MCI Stable_vs_AD | MCI Stable | AD | 0.197 | -0.026 | 0.212 | 0.0013 |
| lhIP1 | MCI Stable_vs_AD | MCI Stable | AD | 0.136 | -0.070 | 0.210 | 0.0010 |
| lhV4 | MCI Stable_vs_AD | MCI Stable | AD | 0.095 | -0.119 | 0.209 | 0.0010 |
| lhs32 | MCI Stable_vs_AD | MCI Stable | AD | 0.143 | -0.058 | 0.208 | 0.0020 |
| rhVMV3 | MCI Stable_vs_AD | MCI Stable | AD | 0.161 | -0.038 | 0.208 | 0.0017 |
| lhOP2-3 | MCI Stable_vs_AD | MCI Stable | AD | 0.142 | -0.049 | 0.205 | 0.0013 |
| rh52 | MCI Stable_vs_AD | MCI Stable | AD | 0.247 | 0.045 | 0.205 | 0.0010 |
| rhFOP5 | MCI Stable_vs_AD | MCI Stable | AD | 0.155 | -0.045 | 0.205 | 0.0023 |
| rhPH | MCI Stable_vs_AD | MCI Stable | AD | 0.212 | 0.015 | 0.204 | 0.0017 |
| lhLO2 | MCI Stable_vs_AD | MCI Stable | AD | 0.037 | -0.164 | 0.204 | 0.0017 |
| rha24pr | MCI Stable_vs_AD | MCI Stable | AD | 0.287 | 0.102 | 0.203 | 0.0020 |
| lhTPOJ1 | MCI Stable_vs_AD | MCI Stable | AD | 0.141 | -0.052 | 0.203 | 0.0023 |
| lh10v | MCI Stable_vs_AD | MCI Stable | AD | 0.229 | 0.026 | 0.203 | 0.0010 |
| lh23d | MCI Stable_vs_AD | MCI Stable | AD | 0.217 | 0.016 | 0.202 | 0.0039 |
| rhMT | MCI Stable_vs_AD | MCI Stable | AD | 0.188 | -0.007 | 0.201 | 0.0020 |
| rhPFt | MCI Stable_vs_AD | MCI Stable | AD | 0.165 | -0.023 | 0.200 | 0.0013 |
| lhFOP5 | MCI Stable_vs_AD | MCI Stable | AD | 0.138 | -0.058 | 0.200 | 0.0020 |
| lhPGs | MCI Stable_vs_AD | MCI Stable | AD | 0.091 | -0.102 | 0.199 | 0.0010 |
| rhMI | MCI Stable_vs_AD | MCI Stable | AD | 0.153 | -0.043 | 0.198 | 0.0023 |
| lhPGi | MCI Stable_vs_AD | MCI Stable | AD | 0.184 | 0.001 | 0.197 | 0.0010 |
| lhOP1 | MCI Stable_vs_AD | MCI Stable | AD | 0.184 | 0.000 | 0.193 | 0.0030 |
| lhPFt | MCI Stable_vs_AD | MCI Stable | AD | 0.185 | 0.000 | 0.192 | 0.0042 |
| rhp24 | MCI Stable_vs_AD | MCI Stable | AD | 0.240 | 0.051 | 0.190 | 0.0036 |
| lhIP2 | MCI Stable_vs_AD | MCI Stable | AD | 0.116 | -0.073 | 0.190 | 0.0033 |
| rh47s | MCI Stable_vs_AD | MCI Stable | AD | 0.171 | -0.013 | 0.188 | 0.0052 |
| lh8Ad | MCI Stable_vs_AD | MCI Stable | AD | 0.205 | 0.024 | 0.186 | 0.0030 |
| lhPH | MCI Stable_vs_AD | MCI Stable | AD | 0.220 | 0.030 | 0.186 | 0.0033 |
| rhFOP4 | MCI Stable_vs_AD | MCI Stable | AD | 0.237 | 0.054 | 0.184 | 0.0054 |
| rhV3CD | MCI Stable_vs_AD | MCI Stable | AD | 0.112 | -0.076 | 0.184 | 0.0042 |
| rhPCV | MCI Stable_vs_AD | MCI Stable | AD | 0.188 | 0.009 | 0.183 | 0.0045 |
| lhPHA3 | MCI Stable_vs_AD | MCI Stable | AD | 0.231 | 0.047 | 0.182 | 0.0033 |
| lh13l | MCI Stable_vs_AD | MCI Stable | AD | 0.189 | 0.018 | 0.182 | 0.0050 |
| lhFOP1 | MCI Stable_vs_AD | MCI Stable | AD | 0.111 | -0.060 | 0.180 | 0.0050 |
| lhV8 | MCI Stable_vs_AD | MCI Stable | AD | 0.144 | -0.032 | 0.180 | 0.0056 |
| rh6r | MCI Stable_vs_AD | MCI Stable | AD | 0.177 | 0.003 | 0.180 | 0.0052 |
| lh9a | MCI Stable_vs_AD | MCI Stable | AD | 0.142 | -0.032 | 0.180 | 0.0050 |
| rhOP4 | MCI Stable_vs_AD | MCI Stable | AD | 0.158 | -0.021 | 0.179 | 0.0056 |
| rhLIPd | MCI Stable_vs_AD | MCI Stable | AD | 0.093 | -0.079 | 0.179 | 0.0054 |
| lh25 | MCI Stable_vs_AD | MCI Stable | AD | 0.164 | -0.007 | 0.178 | 0.0048 |
| rh6ma | MCI Stable_vs_AD | MCI Stable | AD | 0.108 | -0.068 | 0.178 | 0.0071 |
| lh10d | MCI Stable_vs_AD | MCI Stable | AD | 0.138 | -0.039 | 0.178 | 0.0045 |
| rhV4t | MCI Stable_vs_AD | MCI Stable | AD | 0.124 | -0.047 | 0.178 | 0.0065 |
| rhPSL | MCI Stable_vs_AD | MCI Stable | AD | 0.132 | -0.038 | 0.178 | 0.0069 |
| lhIg | MCI Stable_vs_AD | MCI Stable | AD | 0.140 | -0.027 | 0.175 | 0.0069 |
| rhSTSvp | MCI Stable_vs_AD | MCI Stable | AD | 0.194 | 0.031 | 0.175 | 0.0071 |
| rhPFcm | MCI Stable_vs_AD | MCI Stable | AD | 0.132 | -0.040 | 0.175 | 0.0076 |
| rhSTV | MCI Stable_vs_AD | MCI Stable | AD | 0.111 | -0.058 | 0.174 | 0.0068 |
| lhSCEF | MCI Stable_vs_AD | MCI Stable | AD | 0.115 | -0.053 | 0.174 | 0.0054 |
| rh6a | MCI Stable_vs_AD | MCI Stable | AD | 0.199 | 0.029 | 0.174 | 0.0069 |
| rhs32 | MCI Stable_vs_AD | MCI Stable | AD | 0.197 | 0.028 | 0.173 | 0.0061 |
| lh45 | MCI Stable_vs_AD | MCI Stable | AD | 0.214 | 0.054 | 0.172 | 0.0073 |
| lh7Pm | MCI Stable_vs_AD | MCI Stable | AD | 0.087 | -0.081 | 0.171 | 0.0076 |
| lhFST | MCI Stable_vs_AD | MCI Stable | AD | 0.123 | -0.045 | 0.170 | 0.0063 |
| rhLO2 | MCI Stable_vs_AD | MCI Stable | AD | 0.106 | -0.067 | 0.170 | 0.0087 |
| rhMBelt | MCI Stable_vs_AD | MCI Stable | AD | 0.165 | 0.003 | 0.170 | 0.0073 |
| rhLO1 | MCI Stable_vs_AD | MCI Stable | AD | 0.073 | -0.095 | 0.163 | 0.0104 |
| lh9-46d | MCI Stable_vs_AD | MCI Stable | AD | 0.165 | 0.005 | 0.163 | 0.0106 |
| rhIg | MCI Stable_vs_AD | MCI Stable | AD | 0.145 | -0.011 | 0.163 | 0.0104 |
| rh7m | MCI Stable_vs_AD | MCI Stable | AD | 0.154 | -0.007 | 0.161 | 0.0134 |
| rh9-46d | MCI Stable_vs_AD | MCI Stable | AD | 0.096 | -0.059 | 0.160 | 0.0123 |
| rhOP1 | MCI Stable_vs_AD | MCI Stable | AD | 0.127 | -0.029 | 0.158 | 0.0150 |
| lhv23ab | MCI Stable_vs_AD | MCI Stable | AD | 0.105 | -0.049 | 0.156 | 0.0170 |
| lhVMV3 | MCI Stable_vs_AD | MCI Stable | AD | 0.150 | -0.001 | 0.156 | 0.0150 |
| lhSTSva | MCI Stable_vs_AD | MCI Stable | AD | 0.119 | -0.035 | 0.156 | 0.0159 |
| lhRSC | MCI Stable_vs_AD | MCI Stable | AD | 0.103 | -0.056 | 0.155 | 0.0158 |
| rh47m | MCI Stable_vs_AD | MCI Stable | AD | 0.127 | -0.022 | 0.154 | 0.0134 |
| rhLBelt | MCI Stable_vs_AD | MCI Stable | AD | 0.165 | 0.012 | 0.154 | 0.0154 |
| rh23d | MCI Stable_vs_AD | MCI Stable | AD | 0.143 | -0.010 | 0.154 | 0.0134 |
| lhMST | MCI Stable_vs_AD | MCI Stable | AD | 0.061 | -0.093 | 0.154 | 0.0161 |
| rhs6-8 | MCI Stable_vs_AD | MCI Stable | AD | 0.133 | -0.019 | 0.154 | 0.0143 |
| lhA5 | MCI Stable_vs_AD | MCI Stable | AD | 0.168 | 0.020 | 0.152 | 0.0172 |
| lhLO3 | MCI Stable_vs_AD | MCI Stable | AD | 0.063 | -0.093 | 0.151 | 0.0214 |
| lhPoI1 | MCI Stable_vs_AD | MCI Stable | AD | 0.203 | 0.049 | 0.149 | 0.0169 |
| rhV4 | MCI Stable_vs_AD | MCI Stable | AD | 0.129 | -0.023 | 0.147 | 0.0222 |
| rhPGp | MCI Stable_vs_AD | MCI Stable | AD | 0.042 | -0.109 | 0.145 | 0.0223 |
| lhPIT | MCI Stable_vs_AD | MCI Stable | AD | 0.107 | -0.036 | 0.145 | 0.0226 |
| lhPFcm | MCI Stable_vs_AD | MCI Stable | AD | 0.164 | 0.021 | 0.145 | 0.0232 |
| rh8C | MCI Stable_vs_AD | MCI Stable | AD | 0.205 | 0.060 | 0.144 | 0.0272 |
| rh6mp | MCI Stable_vs_AD | MCI Stable | AD | 0.193 | 0.049 | 0.143 | 0.0293 |
| rhPIT | MCI Stable_vs_AD | MCI Stable | AD | 0.154 | 0.010 | 0.143 | 0.0248 |
| lhMI | MCI Stable_vs_AD | MCI Stable | AD | 0.116 | -0.023 | 0.142 | 0.0296 |
| rh9a | MCI Stable_vs_AD | MCI Stable | AD | 0.110 | -0.023 | 0.141 | 0.0263 |
| lhV4t | MCI Stable_vs_AD | MCI Stable | AD | 0.078 | -0.057 | 0.140 | 0.0312 |
| lhMBelt | MCI Stable_vs_AD | MCI Stable | AD | 0.173 | 0.040 | 0.139 | 0.0312 |
| lhIPS1 | MCI Stable_vs_AD | MCI Stable | AD | 0.121 | -0.016 | 0.139 | 0.0307 |
| lhOP4 | MCI Stable_vs_AD | MCI Stable | AD | 0.143 | 0.010 | 0.139 | 0.0339 |
| rhV8 | MCI Stable_vs_AD | MCI Stable | AD | 0.115 | -0.024 | 0.139 | 0.0337 |
| rhFFC | MCI Stable_vs_AD | MCI Stable | AD | 0.151 | 0.017 | 0.139 | 0.0296 |
| lhPCV | MCI Stable_vs_AD | MCI Stable | AD | 0.177 | 0.046 | 0.137 | 0.0356 |
| lh7m | MCI Stable_vs_AD | MCI Stable | AD | 0.138 | 0.008 | 0.136 | 0.0339 |
| lh31pv | MCI Stable_vs_AD | MCI Stable | AD | 0.221 | 0.091 | 0.135 | 0.0334 |
| rhd23ab | MCI Stable_vs_AD | MCI Stable | AD | 0.085 | -0.048 | 0.135 | 0.0339 |
| rhp10p | MCI Stable_vs_AD | MCI Stable | AD | 0.094 | -0.037 | 0.134 | 0.0339 |
| rhRSC | MCI Stable_vs_AD | MCI Stable | AD | 0.072 | -0.061 | 0.132 | 0.0393 |
| lhPBelt | MCI Stable_vs_AD | MCI Stable | AD | 0.192 | 0.068 | 0.131 | 0.0389 |
| rhV3B | MCI Stable_vs_AD | MCI Stable | AD | 0.124 | -0.006 | 0.131 | 0.0434 |
| lhMT | MCI Stable_vs_AD | MCI Stable | AD | 0.140 | 0.009 | 0.130 | 0.0408 |
| rhIFJa | MCI Stable_vs_AD | MCI Stable | AD | 0.102 | -0.026 | 0.130 | 0.0445 |
| rhFOP1 | MCI Stable_vs_AD | MCI Stable | AD | 0.087 | -0.037 | 0.129 | 0.0420 |
| lh44 | MCI Stable_vs_AD | MCI Stable | AD | 0.142 | 0.014 | 0.129 | 0.0481 |
| rhVMV1 | MCI Stable_vs_AD | MCI Stable | AD | 0.115 | -0.008 | 0.128 | 0.0479 |
| lhSTV | MCI Stable_vs_AD | MCI Stable | AD | 0.073 | -0.052 | 0.128 | 0.0484 |
| rhVIP | MCI Stable_vs_AD | MCI Stable | AD | 0.141 | 0.015 | 0.127 | 0.0483 |
| lha10p | MCI Stable_vs_AD | MCI Stable | AD | 0.083 | -0.041 | 0.127 | 0.0447 |
| rh45 | MCI Stable_vs_AD | MCI Stable | AD | 0.153 | 0.032 | 0.127 | 0.0466 |
| lhp9-46v | MCI Stable_vs_AD | MCI Stable | AD | 0.185 | 0.066 | 0.126 | 0.0481 |
| lhTF | MCI Stable_vs_AD | MCI Stable | AD | 0.103 | 0.232 | -0.128 | 0.0481 |
| rh2 | MCI Stable_vs_AD | MCI Stable | AD | 0.115 | 0.252 | -0.136 | 0.0334 |
| lhPeEc | MCI Stable_vs_AD | MCI Stable | AD | 0.041 | 0.183 | -0.139 | 0.0317 |
| lhTGv | MCI Stable_vs_AD | MCI Stable | AD | -0.004 | 0.146 | -0.148 | 0.0197 |
| lh3a | MCI Stable_vs_AD | MCI Stable | AD | 0.122 | 0.271 | -0.154 | 0.0150 |
| rh1 | MCI Stable_vs_AD | MCI Stable | AD | 0.156 | 0.324 | -0.165 | 0.0075 |
| rhPeEc | MCI Stable_vs_AD | MCI Stable | AD | -0.067 | 0.106 | -0.174 | 0.0061 |
| lh1 | MCI Stable_vs_AD | MCI Stable | AD | 0.175 | 0.353 | -0.181 | 0.0063 |
| lh5m | MCI Stable_vs_AD | MCI Stable | AD | 0.064 | 0.245 | -0.186 | 0.0030 |
| lhH | MCI Stable_vs_AD | MCI Stable | AD | -0.292 | 0.006 | -0.190 | 0.0013 |
| rhPreS | MCI Stable_vs_AD | MCI Stable | AD | -0.148 | 0.045 | -0.199 | 0.0020 |
| rhTGd | MCI Stable_vs_AD | MCI Stable | AD | -0.028 | 0.169 | -0.200 | 0.0017 |
| rhV6 | MCI Stable_vs_AD | MCI Stable | AD | 0.003 | 0.203 | -0.201 | 0.0013 |
| rh3a | MCI Stable_vs_AD | MCI Stable | AD | 0.137 | 0.359 | -0.212 | 0.0013 |
| lhPreS | MCI Stable_vs_AD | MCI Stable | AD | -0.144 | 0.093 | -0.240 | 0.0006 |
| lhTE1a | MCI Stable_vs_AD | MCI Stable | AD | 0.012 | 0.265 | -0.256 | 0.0010 |
| lhEC | MCI Stable_vs_AD | MCI Stable | AD | -0.088 | 0.174 | -0.257 | 0.0006 |
| rhH | MCI Stable_vs_AD | MCI Stable | AD | -0.232 | 0.109 | -0.260 | 0.0006 |
| rhEC | MCI Stable_vs_AD | MCI Stable | AD | -0.130 | 0.153 | -0.281 | 0.0006 |
| lhV2 | MCI Stable_vs_AD | MCI Stable | AD | -0.187 | 0.098 | -0.289 | 0.0006 |
| lhTGd | MCI Stable_vs_AD | MCI Stable | AD | -0.087 | 0.230 | -0.310 | 0.0006 |
| rhV2 | MCI Stable_vs_AD | MCI Stable | AD | -0.311 | 0.008 | -0.328 | 0.0006 |
| lhV1 | MCI Stable_vs_AD | MCI Stable | AD | -0.185 | 0.261 | -0.428 | 0.0006 |
| rh3b | MCI Stable_vs_AD | MCI Stable | AD | -0.024 | 0.427 | -0.443 | 0.0006 |
| lh3b | MCI Stable_vs_AD | MCI Stable | AD | -0.020 | 0.436 | -0.457 | 0.0006 |
| rhV1 | MCI Stable_vs_AD | MCI Stable | AD | -0.312 | 0.206 | -0.506 | 0.0006 |
| rhPeEc | MCI Stable_vs_MCI Progressive | MCI Stable | MCI Progressive | -0.067 | 0.194 | -0.258 | 0.0360 |
| rhs32 | MCI Stable_vs_MCI Progressive | MCI Stable | MCI Progressive | 0.197 | -0.039 | 0.252 | 0.0360 |
| rhIPS1 | MCI Stable_vs_MCI Progressive | MCI Stable | MCI Progressive | 0.200 | -0.073 | 0.276 | 0.0360 |
| lhIP0 | MCI Stable_vs_MCI Progressive | MCI Stable | MCI Progressive | 0.108 | -0.194 | 0.321 | 0.0360 |

### 4. **APOE And Normative MIND Analyses**

We conducted ordinary least squares (OLS) regressions to evaluate the effect of APOE genotype on the number of extreme **positive** and **negative** deviations in regional MIND z-scores, using ε3 homozygotes as the reference group and controlling for age and sex. For both outcomes, we also examined interaction terms with age and sex.

The sex composition for the groups were as follows: ε3 homozygotes: N=289 females and N=267 males; ε2 carriers: N=39 females and N=54 males; ε4 heterozygotes: N=283 females and N=234 males; ε4 homozygotes: N=62 females and N=71 males. For the analyses, males were coded as 1 and females as 0.

Supplementary Table 3 shows the comparison of nested models for both positive and negative deviation counts. For positive deviations, the basic additive model provided the best fit (lowest AIC = 11,130.3), with no significant improvement from interaction terms. For negative deviations, the sex interaction model was the best (AIC = 10,397.7), showing significant improvement over the basic model (p = 0.040).

**Supplementary Table 3** – Summary of the comparison of nested APOE models for both positive and negative deviation counts

| **Outcome** | **Model** | **Log-Likelihood** | **AIC** |
| --- | --- | --- | --- |
| Positive Deviation Count | Basic | -5559.14 | 11130.28 |
| Positive Deviation Count | Age Interaction | -5557.69 | 11133.38 |
| Positive Deviation Count | Sex Interaction | -5557.56 | 11133.11 |
| Positive Deviation Count | Full Interaction | -5555.14 | 11142.27 |
| Negative Deviation Count | Basic | -5193.99 | 10399.99 |
| Negative Deviation Count | Age Interaction | -5192.6 | 10403.2 |
| Negative Deviation Count | Sex Interaction | -5189.85 | 10397.7 |
| Negative Deviation Count | Full Interaction | -5183.57 | 10399.13 |

Supplementary Table 4 presents the full regression results for the best-fitting models. For positive deviations, only ε2 carriers showed significant differences from ε3 homozygotes (β = 4.76, p = 0.016). For negative deviations, the key finding was the significant APOE × sex interaction for ε4 homozygotes (β = -7.04, p = 0.006), indicating that the effect of ε4 homozygosity is largely confined to females.

**Supplementary Table 4 –** Full results from the best fitting models assessing APOE genotype effects on cortical deviation burden.

| **Outcome** | **Predictor** | **Coeff.** | **SE** | **t-stat** | **p-val** | **95% CI Lower** | **95% CI Upper** |
| --- | --- | --- | --- | --- | --- | --- | --- |
| Positive Deviation Count (Basic Model) | Intercept | 1.666 | 4.126 | 0.404 | 0.686 | -6.429 | 9.761 |
| Positive Deviation Count (Basic Model) | APOE: e2 carriers | 4.762 | 1.972 | 2.415 | 0.016 | 0.894 | 8.63 |
| Positive Deviation Count (Basic Model) | APOE: e4 heterozygotes | 0.739 | 1.071 | 0.69 | 0.49 | -1.362 | 2.84 |
| Positive Deviation Count (Basic Model) | APOE: e4 homozygotes | -1.261 | 1.703 | -0.741 | 0.459 | -4.601 | 2.079 |
| Positive Deviation Count (Basic Model) | Age | 0.087 | 0.055 | 1.598 | 0.11 | -0.02 | 0.194 |
| Positive Deviation Count (Basic Model) | Sex | 2.517 | 0.975 | 2.582 | 0.01 | 0.604 | 4.43 |
| Negative Deviation Count (Sex Interaction Model) | Intercept | 16.457 | 3.159 | 5.209 | <0.01 | 10.259 | 22.654 |
| Negative Deviation Count (Sex Interaction Model) | APOE: e2 carriers | 1.563 | 2.261 | 0.691 | 0.49 | -2.872 | 5.998 |
| Negative Deviation Count (Sex Interaction Model) | APOE: e4 heterozygotes | 0.236 | 1.104 | 0.214 | 0.831 | -1.931 | 2.403 |
| Negative Deviation Count (Sex Interaction Model) | APOE: e4 homozygotes | 7.485 | 1.86 | 4.024 | <0.01 | 3.836 | 11.133 |
| Negative Deviation Count (Sex Interaction Model) | Sex | -1.843 | 1.12 | -1.645 | 0.1 | -4.041 | 0.355 |
| Negative Deviation Count (Sex Interaction Model) | APOE x Sex: e2 carriers | -2.159 | 2.994 | -0.721 | 0.471 | -8.033 | 3.715 |
| Negative Deviation Count (Sex Interaction Model) | APOE x Sex: e4 heterozygotes | -0.282 | 1.617 | -0.175 | 0.861 | -3.454 | 2.89 |
| Negative Deviation Count (Sex Interaction Model) | APOE x Sex: e4 homozygotes | -7.042 | 2.556 | -2.755 | 0.006 | -12.056 | -2.027 |
| Negative Deviation Count (Sex Interaction Model) | Age | -0.121 | 0.041 | -2.92 | 0.004 | -0.201 | -0.04 |

Supplementary Table 5 reports post-hoc pairwise comparisons using Tukey's HSD test. For positive deviations, only the comparison between ε2 carriers and ε4 homozygotes was significant (p = 0.046). For negative deviations, ε4 homozygotes differed significantly from both ε3 homozygotes (p = 0.009) and ε4 heterozygotes (p = 0.018).

**Supplementary Table 5** - Post-hoc Tukey HSD tests comparing APOE genotype groups on deviation counts.

| **Outcome** | **Group 1** | **Group 2** | **Mean Difference** | **p-value (adj)** | **CI Lower** | **CI Upper** |
| --- | --- | --- | --- | --- | --- | --- |
| Positive Deviation Count | e2 carriers | e3 homozygotes | -4.748 | 0.075 | -9.809 | 0.313 |
| Positive Deviation Count | e2 carriers | e4 heterozygotes | -4.147 | 0.155 | -9.235 | 0.942 |
| Positive Deviation Count | e2 carriers | e4 homozygotes | -6.188 | 0.046 | -12.295 | -0.082 |
| Positive Deviation Count | e3 homozygotes | e4 heterozygotes | 0.601 | 0.944 | -2.159 | 3.361 |
| Positive Deviation Count | e3 homozygotes | e4 homozygotes | -1.44 | 0.831 | -5.801 | 2.921 |
| Positive Deviation Count | e4 heterozygotes | e4 homozygotes | -2.041 | 0.63 | -6.434 | 2.351 |
| Negative Deviation Count | e2 carriers | e3 homozygotes | -0.492 | 0.988 | -4.336 | 3.351 |
| Negative Deviation Count | e2 carriers | e4 heterozygotes | -0.239 | 0.999 | -4.103 | 3.625 |
| Negative Deviation Count | e2 carriers | e4 homozygotes | 3.568 | 0.196 | -1.069 | 8.205 |
| Negative Deviation Count | e3 homozygotes | e4 heterozygotes | 0.254 | 0.99 | -1.842 | 2.35 |
| Negative Deviation Count | e3 homozygotes | e4 homozygotes | 4.06 | 0.009 | 0.749 | 7.372 |
| Negative Deviation Count | e4 heterozygotes | e4 homozygotes | 3.807 | 0.018 | 0.471 | 7.142 |

### 5. **Neurobiology Maps**

**Supplementary Table 6** - Full details of the spin-test correlation analyses (N=5000 spins, Hungarian Method)

| **Feature** | **MCI Stable pval (spin)** | **MCI Progressive pval (spin)** | **AD pval (spin)** |
| --- | --- | --- | --- |
| **Metabolic oxygen rate** | 0.349 | 0.455 | 0.309 |
| **Metabolic glucose rate** | 0.334 | 0.483 | 0.126 |
| **Blood Volume** | 0.355 | 0.422 | 0.925 |
| **Blood Flow** | 0.03 | 0.925 | 0.279 |
| **Evolutionary Expansion** | 0.005 | 0.014 | 0.994 |
| **Developmental Expansion** | 0.791 | 0.626 | 0.746 |
| **Glycolytic Index** | 0.523 | 0.761 | 0.111 |
| **Cortical Thickness** | 0.146 | 0.245 | 0.365 |
| **Cortical Curvature** | 0.384 | 0.374 | 0.5 |
| **AHBA-PC1** | 0.187 | 0.119 | 0.87 |
| **Neurosynth-PC1** | 0.577 | 0.154 | 0.503 |
| **T1/T2 Ratio** | 0.1 | 0.252 | 0.248 |
| **Principal Gradient** | 0.494 | 0.263 | 0.742 |
| **Allometric Scaling** | 0.002 | 0.697 | <0.001 |
| **Geometric Distance** | 0.383 | 0.31 | 0.926 |
| **5HT1a** | 0.586 | 0.065 | 0.493 |
| **5HT1b** | 0.015 | 0.817 | 0.041 |
| **5HT2a** | 0.001 | 0.707 | <0.001 |
| **5HT4** | 0.028 | 0.46 | 0.007 |
| **5HT6** | 0.001 | 0.135 | 0.059 |
| **5HTT** | 0.289 | 0.173 | 0.883 |
| **A4B2** | <0.001 | 0.494 | 0.151 |
| **CB1** | 0.004 | 0.03 | 0.672 |
| **D1** | <0.001 | 0.014 | 0.366 |
| **D2** | 0.33 | 0.046 | 0.918 |
| **DAT** | 0.167 | 0.153 | 0.587 |
| **GABAa** | 0.002 | 0.574 | 0.005 |
| **H3** | <0.001 | 0.314 | 0.248 |
| **M1** | 0.03 | 0.504 | 0.172 |
| **mGluR5** | <0.001 | 0.062 | 0.008 |
| **MOR** | <0.001 | 0.008 | 0.32 |
| **NET** | 0.007 | 0.803 | 0.966 |
| **NMDA** | 0.028 | 0.39 | 0.483 |
| **VAChT** | 0.007 | 0.216 | 0.619 |
| **Layer I** | 0.834 | 0.534 | 0.306 |
| **Layer II** | 0.854 | 0.769 | 0.509 |
| **Layer III** | 0.784 | 0.277 | 0.753 |
| **Layer IV** | 0.079 | 0.872 | 0.432 |
| **Layer V** | 0.378 | 0.289 | 0.414 |
| **Layer VI** | 0.078 | 0.106 | 0.724 |
| **Externo-pyramidisation** | 0.332 | 0.865 | 0.913 |
| **Heteromodal** | 0.131 | 0.468 | 0.145 |
| **Idiotypic** | 0.181 | 0.545 | 0.001 |
| **Paralimbic** | 0.67 | 0.58 | 0.548 |
| **Unimodal** | 0.711 | 0.689 | 0.142 |

### 6. **Postmortem Neuropathology Analyses**

From the full NACC dataset, we identified *N*=558 participants with both MRI data and completed postmortem information. After excluding participants with invalid pathology scores (missing or not possible to score: 8 = Not assessed, - 9 = Missing/unknown, -4 = Not available) and restricting to those with complete data for all covariates, our final analytic sample included *N=*240 participants. The demographic and clinical characteristics of this subset are summarized in Supplementary Table 7 below, with the distribution of ABC scores across diagnoses in Supplementary Figure 8.

**Supplementary Table 7**- Demographic and Clinical Characteristics of the Autopsy Sample (N=240)

| **Characteristic** | **Overall (N=240)** | **ABC 0 (N=20)** | **ABC 1 (N=13)** | **ABC 2 (N=40)** | **ABC 3 (N=167)** |
| --- | --- | --- | --- | --- | --- |
| **Diagnosis, n (%)** |  |  |  |  |  |
| AD | 144 (60.0%) | 5 (25.0%) | 6 (46.2%) | 14 (35.0%) | 119 (71.3%) |
| MCI Progressive | 69 (28.8%) | 6 (30.0%) | 4 (30.8%) | 17 (42.5%) | 42 (25.1%) |
| MCI Stable | 27 (11.2%) | 9 (45.0%) | 3 (23.1%) | 9 (22.5%) | 6 (3.6%) |
| **Age at MRI, mean (SD)** | 76.3 (10.2) | 72.1 (10.5) | 75.8 (9.7) | 75.4 (10.3) | 77.2 (10.0) |
| **Gender, n (%)** |  |  |  |  |  |
| Female | 142 (59.2%) | 12 (60.0%) | 7 (53.8%) | 24 (60.0%) | 99 (59.3%) |
| Male | 98 (40.8%) | 8 (40.0%) | 6 (46.2%) | 16 (40.0%) | 68 (40.7%) |
| **Time to death (years), mean (SD)** | 3.2 (2.5) | 3.5 (2.7) | 3.3 (2.4) | 3.4 (2.6) | 3.1 (2.4) |
| **Negative deviations, mean (SD)** | 9.5 (18.6) | 7.2 (5.4) | 5.3 (4.6) | 6.9 (9.2) | 10.9 (21.5) |
| **Positive deviations, mean (SD)** | 10.2 (12.3) | 10.5 (11.7) | 9.8 (14.0) | 10.3 (11.5) | 10.1 (12.5) |

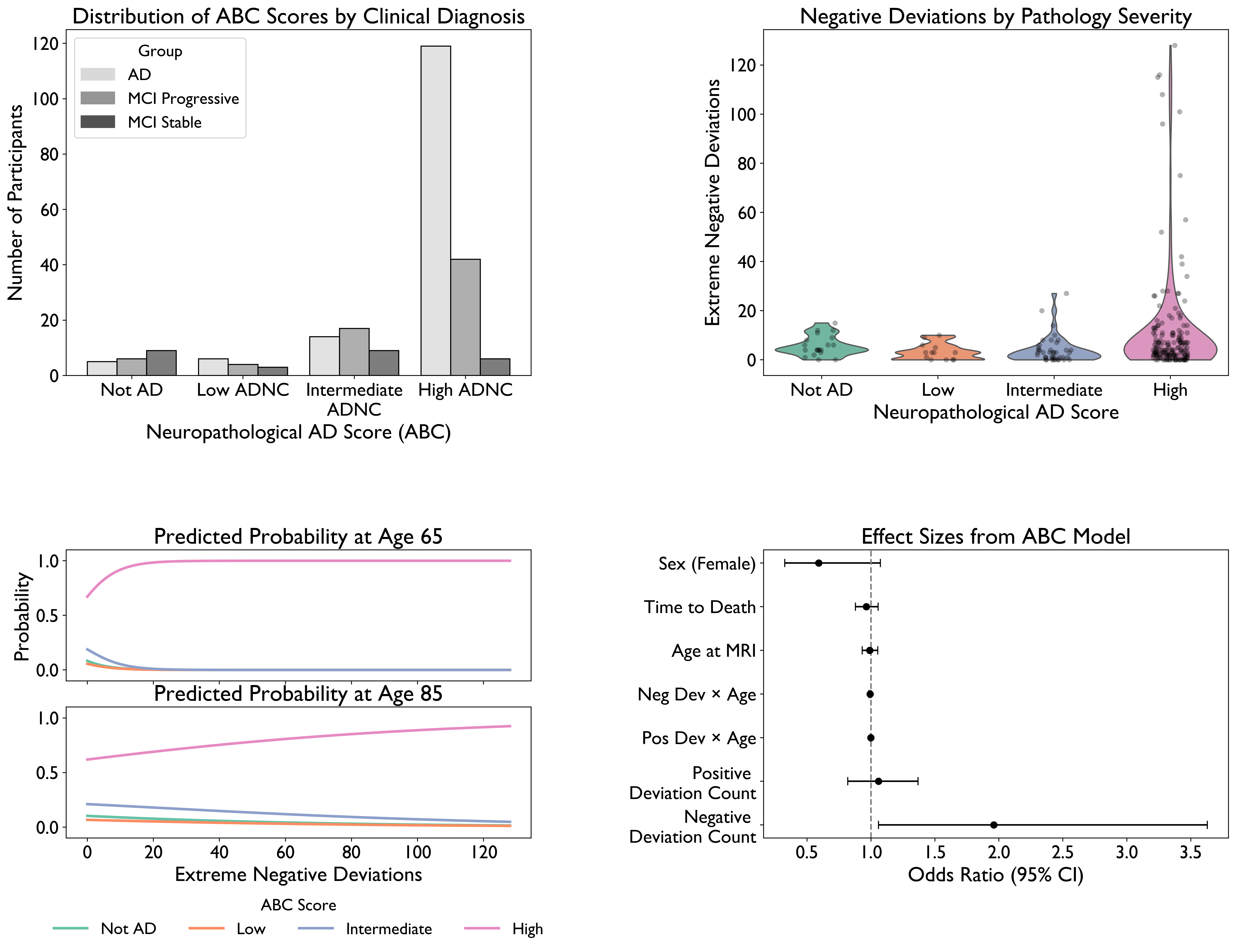

**Supplementary Figure 8**- Histogram showing the distribution of ABC scores (0-3) across diagnostic groups (AD, MCI Progressive, and MCI Stable). Note the higher prevalence of elevated ABC scores (score 3) in the AD group compared to MCI groups.

Neuropathological analyses were performed according to the National Institute on Aging–Alzheimer's Association (NIA-AA) guidelines for the assessment of Alzheimer's disease neuropathologic change (ADNC). All cases underwent standardized autopsy procedures at NACC-affiliated centres. The **ABC** scoring system integrates three key pathological processes:

- **A score (Amyloid-β deposition)**: Based on Thal phase, ranging from 0 (no amyloid) to 5 (widespread distribution)
- **B score (Braak neurofibrillary tangle stage)**: Based on tau pathology distribution, ranging from 0 (none) to 6 (extensive neocortical)
- **C score (CERAD neuritic plaque score)**: Measures neuritic plaque density, ranging from 0 (none) to 3 (frequent)

These individual scores were used to derive the composite ABC score (NPADNC in the NACC dataset), a summary measure of AD pathological change ranging from 0 (not AD) to 3 (high-level AD neuropathologic change).

#### **Statistical Analysis Approach**

We analysed the relationship between structural brain deviations and neuropathological measures using ordinal logistic regression models implemented in the statsmodels Python package (version 0.14.0). To account for the ordinal nature of the ABC score, we used the following model specifications:

##### Basic Model

$$logit(P(ABC\_score \leq k)) = \alphaₖ - (\beta₁ \times neg\_dev\_count + \beta₂ \times pos\_dev\_count + \beta₃ \times age\_at\_MRI + \beta₄ \times sex + \beta₅ \times time\_to\_death)$$

##### Age Interaction Model

$$logit(P(ABC\_score \leq k)) = \alphaₖ - (\beta₁ \times neg\_dev\_count + \beta₂ \times pos\_dev\_count + \beta₃ \times age\_at\_MRI + \beta₄ \times sex + \beta₅ \times time\_to\_death + \beta₆ \times neg\_dev\_count \times age\_at\_MRI + \beta₇ \times pos\_dev\_count \times age\_at\_MRI)$$

##### Time-to-Death Interaction Model

$$logit(P(ABC\_score \leq k)) = \alphaₖ - (\beta₁ \times neg\_dev\_count + \beta₂ \times pos\_dev\_count + \beta₃ \times age\_at\_MRI + \beta₄ \times sex + \beta₅ \times time\_to\_death + \beta₆ \times neg\_dev\_count \times time\_to\_death + \beta₇ \times pos\_dev\_count \times time\_to\_death)$$

Where:

- neg_dev_count and pos_dev_count are the total count of extreme negative and positive deviations (below -1.96 and above 1.96)
- age_at_MRI is the participant's age at MRI acquisition
- sex is coded as 0 (female) or 1 (male)
- time_to_death is the time interval between MRI acquisition and death
- αₖ are the threshold parameters for each level k of the ABC score

Model comparison was performed using the Akaike Information Criterion (AIC), with lower values indicating better relative model fit. The same modelling approach was applied separately to the individual A, B, and C component scores.

#### **Full Model Results for ABC Score Prediction**

**Supplementary Table 8** – Full model results (best model) for the ABC Score Prediction

| A) **Basic Model (AIC = 428.7)** | | | | | |
| --- | --- | --- | --- | --- | --- |
| **Parameter** | **Coefficient** | **Std Error** | **z-value** | **p-value** | **OR [95% CI]** |
| num_extreme_negative_deviations | 0.0536 | 0.026 | 2.056 | 0.040 | 1.06 [1.00-1.11] |
| num_extreme_positive_deviations | 0.0199 | 0.013 | 1.570 | 0.116 | 1.02 [0.99-1.05] |
| age_at_MRI | -0.0539 | 0.019 | -2.841 | 0.004 | 0.95 [0.91-0.98] |
| sex | -0.5456 | 0.302 | -1.806 | 0.071 | 0.58 [0.32-1.05] |
| time_to_death | -0.0534 | 0.046 | -1.160 | 0.246 | 0.95 [0.87-1.04] |
| 0.0/1.0 | -6.7700 | 1.642 | -4.122 | <0.001 | - |
| 1.0/2.0 | -0.5532 | 0.272 | -2.031 | 0.042 | - |
| 2.0/3.0 | 0.0883 | 0.149 | 0.594 | 0.552 | - |

| B) **Age Interaction Model (AIC = 427.8)** | | | | | |
| --- | --- | --- | --- | --- | --- |
| **Parameter** | **Coefficient** | **Std Error** | **z-value** | **p-value** | **OR [95% CI]** |
| num_extreme_negative_deviations | 0.6732 | 0.314 | 2.142 | 0.032 | 1.96 [1.06-3.63] |
| num_extreme_positive_deviations | 0.0566 | 0.131 | 0.432 | 0.666 | 1.06 [0.82-1.37] |
| age_at_MRI | -0.0096 | 0.031 | -0.309 | 0.758 | 0.99 [0.93-1.05] |
| sex | -0.5247 | 0.304 | -1.726 | 0.084 | 0.59 [0.33-1.07] |
| time_to_death | -0.0375 | 0.047 | -0.798 | 0.425 | 0.96 [0.88-1.06] |
| negdev_x_age | -0.0077 | 0.004 | -2.040 | 0.041 | 0.99 [0.98-1.00] |
| posdev_x_age | -0.0005 | 0.002 | -0.271 | 0.786 | 1.00 [1.00-1.00] |
| 0.0/1.0 | -3.1284 | 2.561 | -1.221 | 0.222 | - |
| 1.0/2.0 | -0.5542 | 0.272 | -2.035 | 0.042 | - |
| 2.0/3.0 | 0.0952 | 0.148 | 0.643 | 0.520 | - |

| C) **Time-to-Death Interaction Model (AIC = 429.9)** | | | | | |
| --- | --- | --- | --- | --- | --- |
| **Parameter** | **Coefficient** | **Std Error** | **z-value** | **p-value** | **OR [95% CI]** |
| num_extreme_negative_deviations | 0.1325 | 0.069 | 1.930 | 0.054 | 1.14 [1.00-1.31] |
| num_extreme_positive_deviations | 0.0625 | 0.037 | 1.709 | 0.087 | 1.06 [0.99-1.14] |
| age_at_MRI | -0.0530 | 0.019 | -2.746 | 0.006 | 0.95 [0.91-0.99] |
| sex | -0.5503 | 0.302 | -1.822 | 0.068 | 0.58 [0.32-1.04] |
| time_to_death | 0.0348 | 0.069 | 0.506 | 0.613 | 1.04 [0.90-1.18] |
| negdev_x_ttd | -0.0103 | 0.007 | -1.385 | 0.166 | 0.99 [0.98-1.00] |
| posdev_x_ttd | -0.0053 | 0.004 | -1.295 | 0.195 | 0.99 [0.99-1.00] |
| 0.0/1.0 | -6.0308 | 1.711 | -3.526 | <0.001 | - |
| 1.0/2.0 | -0.5562 | 0.273 | -2.041 | 0.041 | - |
| 2.0/3.0 | 0.0966 | 0.148 | 0.651 | 0.515 | - |

To examine whether the relationship between structural deviation patterns and pathology burden differed by clinical diagnosis, we also stratified our analyses by diagnostic group (AD, MCI Progressive, MCI Stable). The results (Supplementary Table 9) revealed consistent patterns across diagnostic groups, with negative deviations showing positive associations with ABC score in both AD and MCI Progressive groups, although with varying effect sizes. In the AD group, there was a stronger effect of negative deviations (OR = 3.48, 95% CI: 0.75-16.01) and a more pronounced age interaction, suggesting that younger AD patients with high deviation counts may have particularly severe pathology. The MCI Stable group showed an opposite pattern (OR = 0.28), although with wide confidence intervals reflecting the small sample size (N=27).

**Supplementary Table 9** - ABC Models Stratified by Diagnostic Group

##### **A) AD Group (N=144, Model: Age Interaction)**

| Parameter | Coefficient | Std Error | z-value | p-value | OR [95% CI] |
| --- | --- | --- | --- | --- | --- |
| num_extreme_negative_deviations | 1.2478 | 0.780 | 1.599 | 0.110 | 3.48 [0.75-16.01] |
| num_extreme_positive_deviations | 0.0562 | 0.326 | 0.172 | 0.863 | 1.06 [0.56-2.01] |
| age_at_MRI | -0.0565 | 0.062 | -0.916 | 0.360 | 0.95 [0.84-1.07] |
| sex[male] | -0.7897 | 0.572 | -1.381 | 0.167 | 0.45 [0.15-1.39] |
| time_to_death | -0.0340 | 0.088 | -0.389 | 0.697 | 0.97 [0.81-1.15] |
| negdev_x_age | -0.0145 | 0.009 | -1.572 | 0.116 | 0.99 [0.97-1.00] |
| posdev_x_age | -0.0002 | 0.004 | -0.049 | 0.961 | 1.00 [0.99-1.01] |

##### **B) MCI Progressive Group (N=69, Model: Age Interaction)**

| Parameter | Coefficient | Std Error | z-value | p-value | OR [95% CI] |
| --- | --- | --- | --- | --- | --- |
| num_extreme_negative_deviations | 0.5179 | 0.500 | 1.036 | 0.300 | 1.68 [0.63-4.47] |
| num_extreme_positive_deviations | 0.1462 | 0.287 | 0.509 | 0.611 | 1.16 [0.66-2.03] |
| age_at_MRI | 0.0346 | 0.066 | 0.528 | 0.598 | 1.04 [0.91-1.18] |
| sex[male] | -0.2695 | 0.506 | -0.533 | 0.594 | 0.76 [0.28-2.06] |
| time_to_death | 0.1095 | 0.094 | 1.168 | 0.243 | 1.12 [0.93-1.34] |
| negdev_x_age | -0.0062 | 0.006 | -0.988 | 0.323 | 0.99 [0.98-1.01] |
| posdev_x_age | -0.0022 | 0.004 | -0.562 | 0.574 | 1.00 [0.99-1.01] |

##### **C) MCI Stable Group (N=27, Model: Age Interaction)**

| **Parameter** | **Coefficient** | **Std Error** | **z-value** | **p-value** | **OR [95% CI]** |
| --- | --- | --- | --- | --- | --- |
| num_extreme_negative_deviations | -1.2809 | 1.464 | -0.875 | 0.382 | 0.28 [0.02-4.93] |
| num_extreme_positive_deviations | 0.7670 | 0.967 | 0.793 | 0.428 | 2.15 [0.32-14.33] |
| age_at_MRI | 0.0311 | 0.114 | 0.272 | 0.786 | 1.03 [0.82-1.29] |
| sex[male] | -0.8382 | 0.931 | -0.901 | 0.368 | 0.43 [0.07-2.68] |
| time_to_death | -0.1304 | 0.112 | -1.163 | 0.245 | 0.88 [0.70-1.09] |
| negdev_x_age | 0.0137 | 0.017 | 0.812 | 0.417 | 1.01 [0.98-1.05] |
| posdev_x_age | -0.0104 | 0.012 | -0.849 | 0.396 | 0.99 [0.97-1.01] |

#### **Results from Models of Individual A, B, C Components**

To investigate which aspects of AD pathology were most strongly associated with structural deviations, we examined models for each component of the ABC score (Supplementary Table 10). The B score (Braak stage) showed the strongest association with negative deviations, particularly when accounting for age interactions. This suggests that the tau-related aspects of AD pathology may be more closely linked to structural network disruptions than amyloid pathology. The C score (neuritic plaque density) showed minimal association with deviation counts, suggesting that structural deviation patterns may not be strongly related to this aspect of AD pathology.

**Supplementary Table 10 –** Results from models of individual A, B and C Components

##### **A) A Score Model (AIC = 593.6)**

| **Parameter** | **Coefficient** | **Std Error** | **z-value** | **p-value** | **OR [95% CI]** |
| --- | --- | --- | --- | --- | --- |
| num_extreme_negative_deviations | 0.0111 | 0.009 | 1.272 | 0.204 | 1.01 [0.99-1.03] |
| num_extreme_positive_deviations | 0.0031 | 0.007 | 0.422 | 0.673 | 1.00 [0.99-1.02] |
| age_at_MRI | -0.0442 | 0.016 | -2.805 | 0.005 | 0.96 [0.93-0.99] |
| sex[male] | -0.6911 | 0.268 | -2.580 | 0.010 | 0.50 [0.30-0.85] |
| time_to_death | -0.0544 | 0.041 | -1.317 | 0.188 | 0.95 [0.87-1.03] |

##### **B) B Score Model with Age Interaction (AIC = 651.8)**

| **Parameter** | **Coefficient** | **Std Error** | **z-value** | **p-value** | **OR [95% CI]** |
| --- | --- | --- | --- | --- | --- |
| num_extreme_negative_deviations | 0.2534 | 0.196 | 1.294 | 0.196 | 1.29 [0.88-1.89] |
| num_extreme_positive_deviations | -0.1087 | 0.077 | -1.408 | 0.159 | 0.90 [0.77-1.04] |
| age_at_MRI | -0.0539 | 0.026 | -2.110 | 0.035 | 0.95 [0.90-1.00] |
| sex[male] | -0.4626 | 0.268 | -1.728 | 0.084 | 0.63 [0.37-1.06] |
| time_to_death | 0.0207 | 0.042 | 0.494 | 0.621 | 1.02 [0.94-1.11] |
| negdev_x_age | -0.0027 | 0.002 | -1.100 | 0.271 | 1.00 [0.99-1.00] |
| posdev_x_age | 0.0016 | 0.001 | 1.495 | 0.135 | 1.00 [1.00-1.00] |

##### **C) C Score Model (AIC = 489.3)**

| **Parameter** | **Coefficient** | **Std Error** | **z-value** | **p-value** | **OR [95% CI]** |
| --- | --- | --- | --- | --- | --- |
| num_extreme_negative_deviations | -0.0024 | 0.007 | -0.362 | 0.718 | 1.00 [0.99-1.01] |
| num_extreme_positive_deviations | -0.0005 | 0.008 | -0.066 | 0.947 | 1.00 [0.98-1.02] |
| age_at_MRI | -0.0593 | 0.017 | -3.437 | 0.001 | 0.94 [0.91-0.97] |
| sex[male] | -0.7757 | 0.285 | -2.723 | 0.006 | 0.46 [0.26-0.80] |
| time_to_death | -0.0508 | 0.043 | -1.188 | 0.235 | 0.95 [0.87-1.03] |
